# Multiple sclerosis iPSC derived-pericytes contract poorly but respond robustly to lesion-relevant environmental stimuli

**DOI:** 10.1101/2025.07.15.25331537

**Authors:** Alastair J Fortune, Natalie E King, Ariane Gélinas-Marion, Roisin A Moloney, Jake M Cashion, Kathryn P Burdon, Bruce V Taylor, Brad A Sutherland, Jessica L Fletcher, Nicholas B Blackburn, Kaylene M Young

## Abstract

Multiple sclerosis (MS) is associated with vascular abnormalities including blood-brain-barrier disruption, neurovascular uncoupling and cerebral hypoperfusion. As pericytes regulate vascular integrity and capillary diameter, pericyte dysfunction may contribute to MS pathophysiology. To investigate this, we generated induced pluripotent stem cell-derived pericytes (iPericytes) from individuals with or without relapsing-onset MS (RoMS). RoMS iPericytes exhibited intrinsic, cell-autonomous alterations in gene expression, were larger, and responded poorly to the vasoconstrictor, endothelin-1. Despite intrinsic differences, Control and RoMS iPericytes phagocytosed myelin debris and responded robustly to lesion-relevant stimuli. Hypoxia induced iPericyte proliferation, elongation and angiogenic and immune-related gene expression, while exposure to MS-relevant cytokines (TNFα and IFNγ) exacerbated the endothelin-1-evoked contraction. Both RoMS and cytokine exposure altered the expression of calcium and myosin signalling genes downstream of endothelin-1, including the key contractile regulator *MYLK*. These data support intrinsic pericyte dysfunction and signalling within MS-lesions as drivers of hypoperfusion and neurodegeneration in MS.

## Main

Multiple sclerosis (MS) is characterised by central nervous system (CNS) inflammation, demyelination and neuron death ^1^. In addition to these hallmark pathologies, people with MS experience blood-brain-barrier (BBB) disruption ^2–5^, impaired neurovascular coupling (Marshall et al., 2016; Marshall et al., 2014), and cerebral hypoperfusion in the grey ^6–10^ and white matter ^11–13^. Cerebral hypoperfusion likely drives the development of hypoxia in MS brain tissue ^14–19^, which would exacerbate neuroinflammation and neurodegeneration, and impede remyelination ^20^. It remains unclear however, whether hypoperfusion is a primary pathology in people with MS. Over 200 MS susceptibility variants have been identified ^21^, and many of the genes linked to these variants are expressed by cells within the neurovascular unit, implicating vascular cells as contributing to disease risk ^21,22^. These or other variants could intrinsically change the structure or function of key neurovascular cell types ^23,24^.

The neurovascular unit, composed of endothelial cells, astrocytes, neurons and pericytes, regulates BBB permeability, and modulates blood flow in response to the changing metabolic and oxygen demands of the CNS ^25–27^. Throughout the brain capillary network, a single layer of endothelial cells forms the vessel walls and pericytes, embedded in the basement membrane, extend processes around the vessels that contract and relax to regulate vascular tone ^25,28^. Pericytes can be identified by their expression of platelet-derived growth factor receptor β (PDGFRβ), the NG2 proteoglycan, CD13 and, specifically within the CNS, by ATP13A5 ^29^, and while PDGFRβ^+^ cells are present within MS lesions, the exact identity, functional state or contribution of these cells to MS pathophysiology has not been determined^27,30,31^.

Pericytes perform a diverse range of roles that could enable them to play a primary and / or secondary role in MS pathophysiology ^32,33^. For example, pericytes can produce inflammatory cytokines and enhance macrophage extravasation ^30^. Interestingly, pericyte loss also promotes vascular and immune pathologies, with pericyte depletion being sufficient to reduce BBB integrity, upregulate the expression of leukocyte adhesion molecules on endothelial cells, and promote peripheral immune cell extravasation in the CNS ^34–36^. While pericyte density is reduced in the cerebellum of mice with experimental autoimmune encephalomyelitis (EAE) ^30^, it is unclear if this pericyte loss drives the associated diminished cerebrovascular reactivity or increased leukocyte stalling and extravasation ^37^.

These mouse studies highlight the capacity for pericytes to contribute to MS pathophysiology. However, they cannot determine whether pericytes experience intrinsic dysfunction prior to MS onset, that could also modify their response to pathological signalling within MS lesions. These questions can be answered by differentiating induced pluripotent stem cells (iPSCs) from people with and without relapsing onset MS (RoMS) to obtain pericyte-like cells (iPericytes; ^23,38,39^ for direct interrogation.

Herein, we report that the expression of contractility-related genes is altered in RoMS iPericytes, and that their contractile response to endothelin-1 (ET-1) is impaired. This intrinsic pericyte dysfunction would compromise capillary blood flow regulation and contribute to cerebral hypoperfusion. We also report that pericyte function is modulated by MS lesion-associated stimuli including exposure to myelin debris, hypoxia, and inflammatory cytokines. Notably, pre-exposure to TNFα and IFNγ drives an excessive contractile response to ET-1 that would be predicted to significantly reduce capillary blood flow within lesions. These findings position pericytes as active contributors to MS pathogenesis and progression, and a potential therapeutic target for neuroprotection in MS.

## Results

### Transcriptomic profiling reveals intrinsic alterations in contractile and vascular regulatory pathways in RoMS iPericytes

To determine whether pericytes are intrinsically altered in people with MS, we obtained blood from people without MS (n=10, Controls), MS-affected members of a multi-case family (n=4, Family RoMS cases spanning 2 generations) or unrelated people with RoMS (n=8, RoMS cases). Peripheral blood mononuclear cells were isolated and used for Sendai-virus mediated iPSC generation as previously described ^40,41^. The resulting 22 iPSC lines (**Table 1**) were differentiated into iPericytes using a validated protocol ^38,39^ (**Fig. 1a**), and immunocytochemistry performed to confirm that > 95% of the cells in each culture expressed the pericyte proteins PDGFRβ and CD13 (*ANPEP*) (**Fig. 1b**; **Fig. S1a**). RNA was subsequently isolated from iPericytes and bulk RNA sequencing performed to confirm successful differentiation. iPericytes exhibited a transcriptional profile consistent with human brain vascular pericytes, with a high level of expression of pericyte genes and minimal expression of genes specific to other CNS-resident cell types (**Fig. S1b**). Of the 551 genes potentially impacted by MS-associated risk variants ^22^, 427 were expressed by iPericytes, with 142 being upregulated during iPericyte differentiation (**Fig. S1c**). These data indicate that any dysregulation of these genes that increases MS risk could be impacting pericyte function.

**Figure 1:**
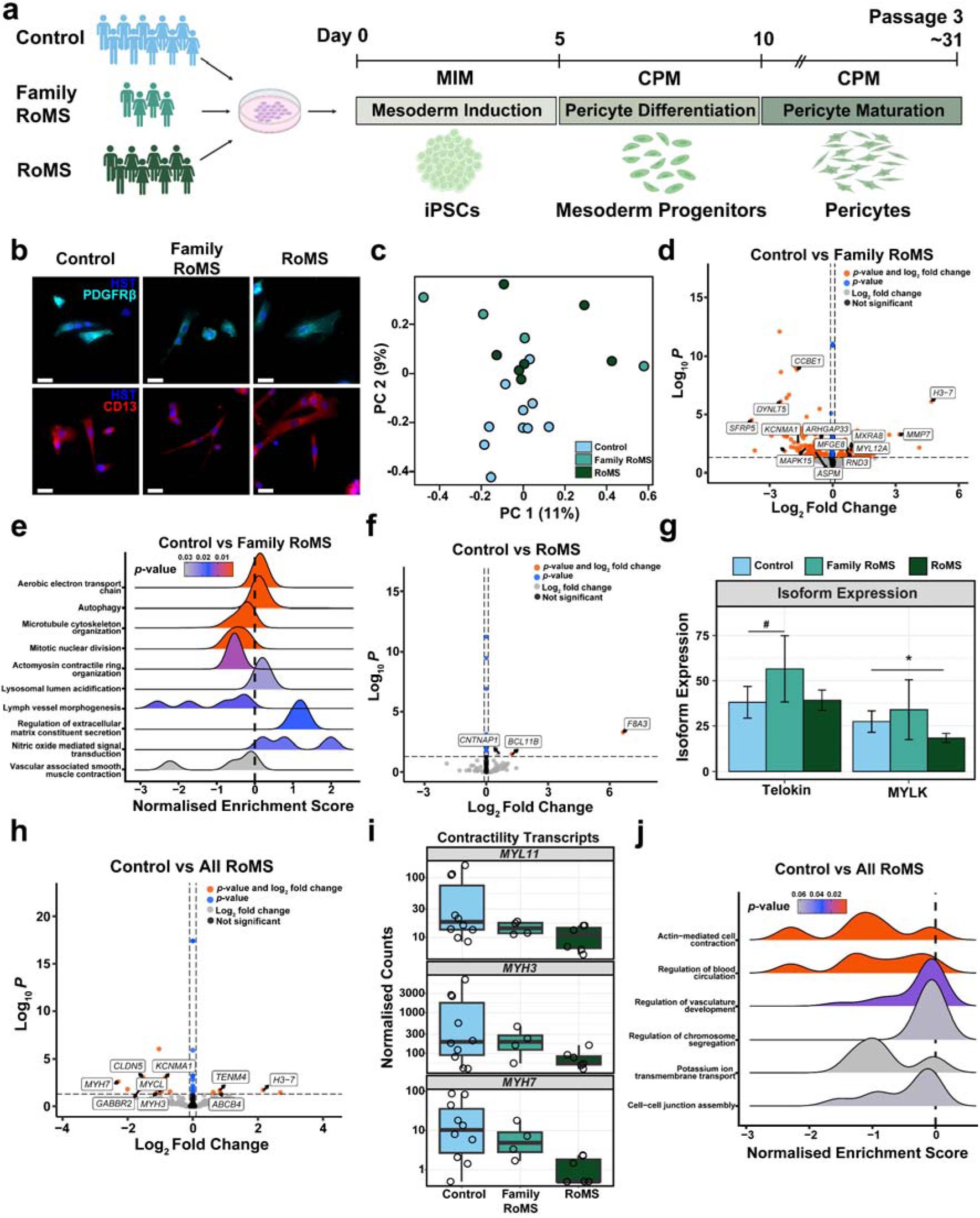
Gene expression differs between iPericytes from people with or without RoMS. (a) Experimental design and timeline of pericyte differentiation. (b) Images (20x) of PDGFRβ (cyan) and CD13 (red) expression by iPericytes. Scale bars, 50µm. (c) Principal component analysis of gene expression incorporating 1 factor of unwanted variation (n=10 Control, n=4 Family RoMS and n=7 RoMS cell lines). (d) Volcano plot showing genes that are differentially expressed between Control (n=10 lines) and Family RoMS (n=4 lines) iPericytes. Adjusted p-value <0.05 and Log2 fold change ≥0.1. (e) Ridge plot showing gene set enrichment analysis results comparing Control and Family RoMS iPericytes. (f) Volcano plot showing genes that are differentially expressed between Control (n=10 lines) and RoMS (n=7 lines) iPericytes. (g) Barplots showing the isoform expression for *MYLK*. Data expressed as mean ± standard deviation for n=10 Control or n=4 Family RoMS or n=7 RoMS iPericyte cultures. (h) Volcano plot showing genes that are differentially expressed between Control (n=10 lines) and Family RoMS (n=4 lines) combined with RoMS (n=7 lines) iPericytes. (i) Boxplots showing gene expression in Control, Family RoMS and RoMS iPericytes, for DEGs with known biological roles that may impact pericyte contractility. Data expressed as median ± interquartile range. Dot points represent individual cultures from n=10 Control or n=4 Family RoMS or n=7 RoMS iPericyte cultures. (j) Ridge plot showing gene set enrichment analysis results comparing Control and all RoMS iPericytes. False discovery rate adjustment was used for multiple testing for RNAseq data.

**Table 1:** iPSC line details and disease status.

| **hPSC<sup>reg</sup>® Line** | **Group** | **Sex** | **Age at Collection** |
| --- | --- | --- | --- |
| MNZTASi009-A | Control | F |  |
| MNZTASi019-A | Control | F |  |
| MNZTASi020-A | Control | F |  |
| MNZTASi021-A | Control | M |  |
| MNZTASi022-A | Control | F |  |
| MNZTASi024-A | Control | F |  |
| MNZTASi032-A | Control | M |  |
| MNZTASi040-A | Control | M |  |
| MNZTASi050-A | Control | M |  |
| MNZTASi070-A | Control | M |  |
| MNZTASi002-A | Family RoMS | F |  |
| MNZTASi003-A | Family RoMS | F |  |
| MNZTASi004-A | Family RoMS | F |  |
| MNZTASi005-A | Family RoMS | M |  |
| MNZTASi007-A | RoMS | F |  |
| MNZTASi010-A | RoMS | F |  |
| MNZTASi014-A | RoMS | F |  |
| MNZTASi016-A | RoMS | F |  |
| MNZTASi017-A | RoMS | F |  |
| MNZTASi018-A | RoMS | F |  |
| MNZTASi033-A | RoMS | M |  |
| MNZTASi037-A | RoMS | M |  |

To determine whether RoMS iPericytes have transcriptional signatures distinct from Controls, we compared the transcriptome of Control, Family RoMS, and RoMS iPericytes. Global gene expression analysis using PCA revealed partial separation along PC2 (9% of variance) of Control, Family RoMS, and RoMS samples (**Fig. 1c**). Our initial differential gene expression analysis compared Control and Family RoMS iPericytes (log_₂_FC>0.1, α<0.05) and identified 251 differentially expressed genes (DEGs) (**Fig. 1d**), including regulators of cell contraction, such as decreased expression of *myosin light chain 12A (MYL12A;* LFC=0.3, Padj=0.048) and *calcium-activated potassium channel subunit* α*1* (*KCNMA1*; LFC=-1.7, Padj=0.003) (**Data TS1**) ^42,43^. Gene set enrichment analysis revealed that vascular smooth muscle contraction and microtubule organisation pathways were suppressed (**Fig. 1e, Data TS2**), suggesting that contractility may be disturbed in Family RoMS iPericytes. As alternative splicing of genes can impact elements of the contractile machinery in pericytes ^44^, we also performed differential transcript usage (DTU) analysis.

Comparing Control and Family RoMS iPericytes we identified 18 genes with DTU (**Fig. S1d, Data TS3**), including *RND3* (a gene associated with an MS risk variant ^21^), which encodes a Rho GTPase that can regulate contractility and blood flow ^45,46^. In Family RoMS iPericytes, we also found that the *CDKN2A* isoform encoding the p16^INK4A^ cellular senescence protein was upregulated (**Fig. S1e**). As p16^INK4A^ expression is upregulated in neural progenitor cells in primary progressive MS postmortem brain tissue and in iPSC-derived neural progenitor cells from progressive MS donors ^47,48^, this may represent a common phenotype across cell types.

Our comparison of Control and RoMS iPericyte samples identified only 3 differentially expressed genes (**Fig. 1f, Data TS4**). RoMS iPericytes having fewer significant transcriptional changes than Family RoMS iPericytes would be consistent with the cause and level of intrinsic dysfunction being more varied in unrelated MS cases. However, there were also 11 genes with significant DTU (**Fig. S1f, Data TS5**). This included alternative splicing of *myosin light chain kinase* (*MYLK;* Padj=0.015), which was driven by the decreased expression of the canonical *MYLK* transcript (**Fig. 1g, Fig. S1g**), which encodes the calcium-dependent myosin light chain kinase (MLCK). Pericyte contractile state is strongly influenced by the activity of MLCK, which phosphorylates the myosin light chains to drive contraction ^26^, and works in concert with the myosin light chain phosphatase (MLCP), which promotes relaxation (Wilhelm et al., 2025). Interestingly, Family RoMS exhibited increased expression of the shortest characterised isoform of *MYLK*, which encodes the protein Telokin (**Fig. 1g**). Telokin inhibits MLCK-mediated phosphorylation of myosin light chains, while enhancing MLCP activity, thereby desensitizing cells to calcium-mediated contractility ^49,50^. Therefore, while mediated by distinct isoform changes in Family RoMS and RoMS iPericytes, both gene expression changes would diminish MLCK activity and be predicted to reduce pericyte contractility.

To gain further insight into key dysregulated pathways, we compared Control samples with All RoMS (n=12, Family RoMS+RoMS) and identified 18 DEGs (**Fig. 1h, Data TS6**). Downregulated genes included components of the myosin light (*MYL11*) and heavy chains (*MYH3*, *MYH7*) (**Fig. 1i**), and enrichment analysis confirmed that pathways associated with contraction, blood circulation, and vascular development were significantly altered (**Fig. 1j, Data TS7**). These data indicate that Family RoMS and RoMS iPericytes have intrinsic transcriptional changes that are likely to impact actomyosin-mediated contraction.

### RoMS iPericytes are enlarged and contract poorly following endothelin-1 exposure

To determine whether intrinsic changes in RoMS iPericyte gene expression altered iPericyte structure or function, we collected phase contrast images of Control, Family RoMS and RoMS iPericyte cultures and performed a morphological analysis. Across all groups, individual cells adopted a mixture of pericyte standard, spindle, sheet or circular morphologies ^39,51^ (**Fig. S2a**). Standard morphology predominated, and the overall distribution of morphologies was comparable between Control and Family RoMS cultures (**Fig. S2a**). Consistent with this, iPericyte elongation, indicated by maximum Feret diameter (diameter at widest point / perimeter length), was unaffected by MS status (**Fig. S2b**). However, by tracing the perimeter of individual pericytes in each culture (**Fig. 2a**), and quantifying cell area and perimeter length, we determined that Family RoMS and RoMS iPericytes were significantly larger than Control iPericytes (**Fig. 2b,c**). This may be directly due to changes to myosin subunits ^51,52^, but could also be impacted by a change in proliferation kinetics. To evaluate this possibility, Control, Family RoMS and RoMS iPericytes were exposed to the thymidine analogue, ethynyl-2’-deoxyuridine (EdU) for 20 hours. EdU^+^ pericytes were detected in all cultures (**Fig. S2c**), but the proportion of iPericyte that had undergone division and incorporated EdU was unaffected by MS status (**Fig. S2d**). The increased cell size was also unrelated to a change in basal motility (**Fig. S2e-g**) but could reflect a change in contractile state.

**Figure 2:**
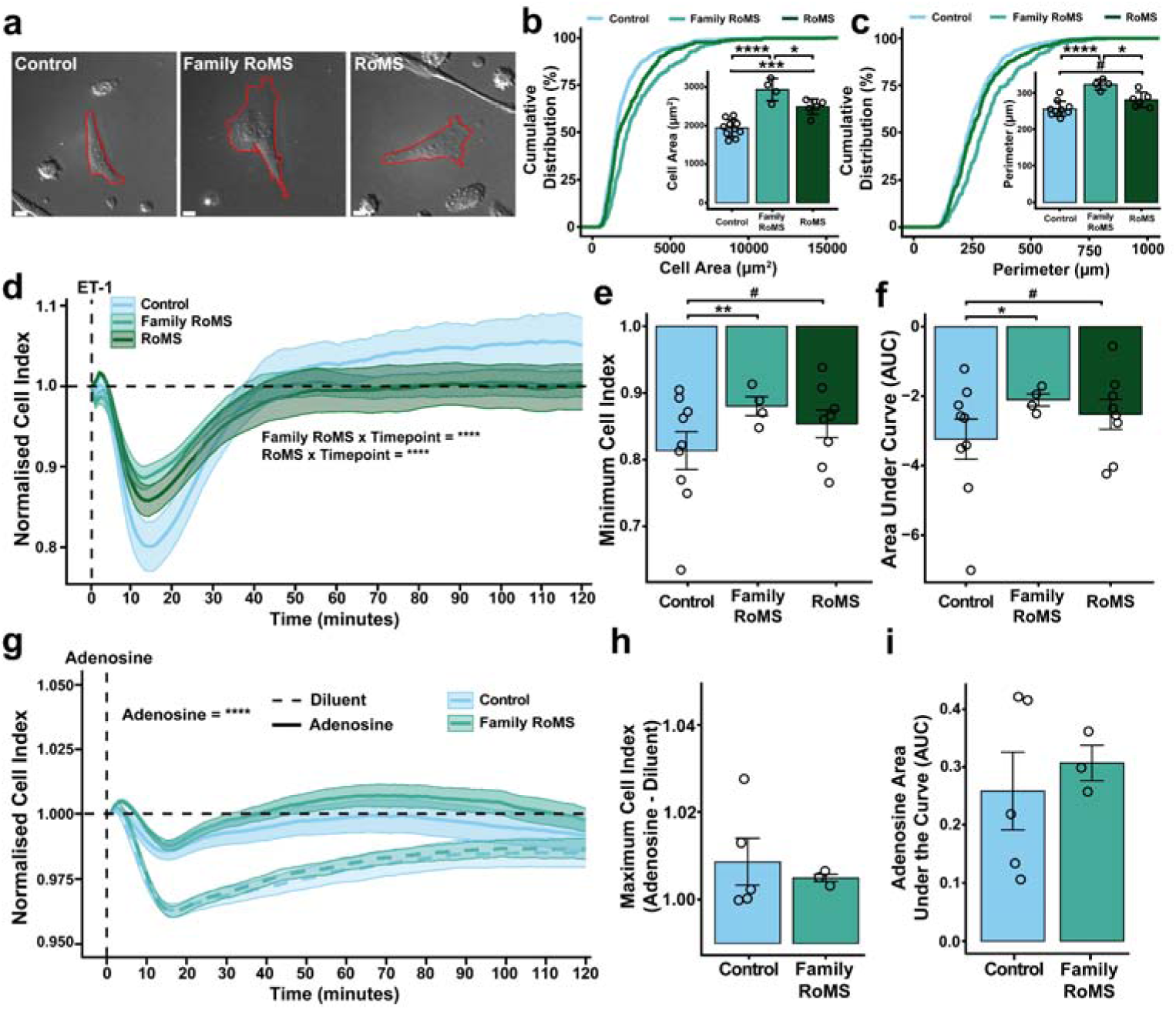
Enlarged RoMS iPericytes have a reduced contractile response to endothelin-1 (ET-1). (a) Images of Control, Family RoMS and RoMS pericytes with their perimeter traced for morphological assessment (red). Scale bars, 10µm. (b) Cumulative distribution plot of iPericyte cell area. > 50 cells evaluated per cell line for n=10 Control, n=4 Family RoMS and n=8 RoMS lines. Inset graph depicts the mean cell area per group ± standard deviation, and dots represent the mean for individual cell lines. (c) Cumulative distribution plot of iPericyte perimeter. > 50 cells evaluated in each of n=10 Control, n=4 Family RoMS and n=8 RoMS lines. Inset graph depicts the mean cell perimeter per group ± standard deviation. (d) Normalised electrical impedance cell index for Control, Family RoMS and RoMS iPericytes responding to ET-1 over a 2 hour period. 2-4 wells evaluated per cell line to obtain the average per cell line. Data presented represent the group mean ± SEM for n=9 Control, n=4 Family RoMS and n=8 RoMS cell lines. (e,f) Quantification of the minimum cell index (maximum pericyte contraction) or area under the curve for the first 20 minutes following ET-1 treatment of Control, Family RoMS and RoMS iPericytes. 2-4 wells were evaluated to obtain the average for each cell line and data are presented as mean for each group ± SEM from n=9 Control, n=4 Family RoMS and n=8 RoMS lines. (g) Normalised electrical impedance cell index for Control and Family RoMS iPericytes following adenosine exposure. 2-4 wells were evaluated to obtain a per cell line average. Data are presented as mean per group ± SEM for n=5 Control or n=3 Family RoMS lines. (h) Quantification of the maximum cell index (level of relaxation) for Control and Family RoMS iPericytes exposed to diluent or adenosine. 2-4 wells were evaluated to obtain a per cell line average. Data presented represent the mean per treatment ± SEM for n=5 Control and n=3 Family RoMS lines. (i) Area under the curve for the first 20 minutes following adenosine treatment of Control or Family RoMS iPericytes. Data presented represent the mean per treatment ± SEM for n=5 Control and n=3 Family RoMS lines. #<0.1, _∗_p<0.05, _∗∗_p<0.01 _∗∗∗_p<0.001, _∗∗∗∗_p<0.0001. Restricted maximum likelihood linear mixed model with pairwise comparisons using Tukey’s adjustment for p-values.

To determine whether RoMS iPericyte contractility was altered, we exposed Control Family RoMS and RoMS iPericytes to endothelin-1 (ET-1). ET-1 is a potent pericyte vasoconstrictor ^39,53,54^ (**Fig. S2h, i**), and iPericytes from all groups express the *EDNRA* transcript, which encodes the primary ET-1 receptor (LFC=-0.00002, padj=0.999; **Fig. S2j**). Following ET-1 application, we quantified the size of the evoked contraction by measuring electrical impedance: when iPericytes contract they reduce their surface area, reducing electrical impedance (cell index) across the plate (**Fig. 2d**). We found that iPericytes from Control, Family RoMS, and RoMS participants had similar contraction kinetics, reaching their maximal contraction after ∼14 minutes. However, the magnitude of contraction differed significantly between groups. The contractile response was significantly impaired in Family RoMS and RoMS iPericyte, with the evoked contractions reaching 64.1% and 78.3% of the maximal Control contraction, respectively (**Fig. 2e**). Furthermore, the volume of contraction, given by the area under the curve (AUC), was markedly decreased in Family RoMS and RoMS iPericytes (**Fig. 2f**). These data demonstrate that the intrinsic gene expression changes within contractile signalling pathways manifests as significantly impaired contraction by Family RoMS and RoMS iPericytes.

As pericytes modulate blood flow through vasoconstriction and vasodilation ^25,39^, we next evaluated the capacity of iPericytes to relax when exposed to the vasodilator, adenosine ^39,51^. iPericyte cultures contracted when the E-plate was removed from the incubator but relaxed significantly following the addition of 10µM adenosine (**Fig. 2g**). We found that Family RoMS iPericytes had equivalent maximum and volume of Control iPericytes, indicating a preserved ability to relax (**Fig. 2h,i**). Pericytes exist in a partially constricted state at rest to provide basal tone for capillaries, but dynamically respond to vasoconstrictors and vasodilators to direct blood flow to metabolically active regions ^25,26^. Within the CNS, a local reduction in pericyte contraction in response to ET-1 would increase blood flow through the capillary bed, however, as we predict this would instead impact tone across the entire RoMS brain capillary network, it would be more likely to increase capillary blood volume but reduce flow rate, leading to hypoperfusion (**Fig. S3a**). Consistent with this hypothesis, hypoperfusion is a feature of clinically isolated syndrome ^55^, a clinical presentation that can precede an MS diagnosis.

### iPericytes phagocytose myelin debris

Intrinsic pericyte dysfunction would be expected to impact vascular function before and after MS onset. However, once MS lesions form, pericytes are exposed to the complex lesion microenvironment. To determine how pericytes might respond to such stimuli, we examined the response of Control and Family RoMS iPericytes to myelin debris, hypoxia and inflammatory cytokines.

Myelin debris present within MS lesions can be phagocytosed and cleared by microglia and astrocytes ^56^, and there is evidence that endothelial cells can phagocytose myelin debris following spinal cord injury or EAE (Zhou et al., 2019a). While pericytes phagocytose aggregated amyloid β42 ^57^, it is unknown whether they support myelin clearance. To evaluate the capacity of iPericytes to phagocytose myelin and determine whether this is influenced by RoMS status, Control and Family RoMS iPericytes were plated onto pHrodo-myelin coated plates and imaged for 24 hours (**Fig. 3a**). pHrodo-myelin was detected inside iPericytes within 20 min, reflecting myelin debris uptake into acidic phagolysosomes (**Fig. 3b**). A loss of pericyte phagocytosis may partly explain why myelin debris clearance is impaired in pericyte-deficient mice following middle cerebral artery occlusion (Shibahara et al., 2020). At 24 hours, >85% of Control and Family RoMS iPericytes were pHrodo-myelin^+^ and the rate of cell labelling was not impacted by RoMS status (**Fig. 3c**). Furthermore, the proportion of iPericyte area with pHrodo^+^ pixels was equivalent in Control and Family RoMS iPericytes (**Fig. 3d**), indicating that Family RoMS iPericytes have a similar capacity to support myelin clearance. Importantly, Control and RoMS iPericytes maintained their normal morphology, and remained proliferative and motile after 20 hours of myelin debris exposure (**Fig. 3e-i**), indicating that myelin phagocytosis is not toxic to iPericytes.

**Figure 3:**
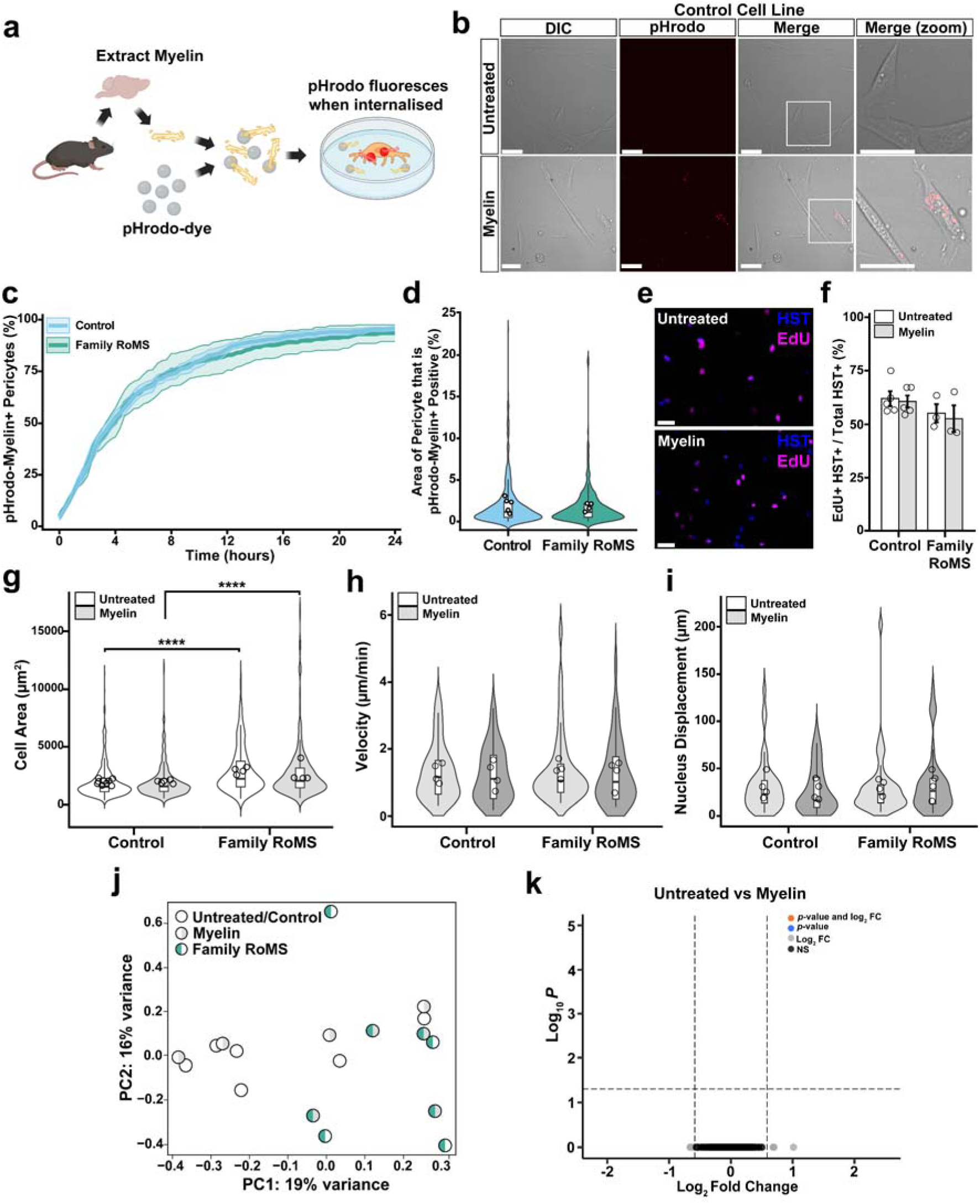
iPericytes phagocytose myelin debris but this is not associated with morphologically or transcriptional changes. (a) Schematic of mouse myelin extraction and the treatment of iPericytes. (b) Epifluorescence and phase contrast images (20x) of iPericytes cultured without (untreated) or with pHrodo-tagged myelin debris for 20 hours. Scale bars, 50µm. (c) Quantification of the percentage (%) of Control or Family RoMS iPericytes that phagocytosed myelin and became pHrodo^+^ over time (imaged every 20 minutes for 20 hours). > 50 cells evaluated per cell line; n=5 Control and n=4 Family RoMS cell lines. Data expressed as mean per group ± SEM. (d) Quantification of iPericytes area with myelin-pHrodo^+^ pixels (%) was calculated for n=5 Control and n=4 Family RoMS lines (> 100 cells evaluated per line). Data expressed as median ± interquartile range, dots represent cell line mean. (e) Images (10x) of iPericytes that incorporated EdU (red) under untreated or myelin conditions. Scale bars, 50µm. (f) The percentage (%) of iPericytes per culture that became EdU+ over 20 hours in untreated and myelin conditions. 4-9 wells were evaluated per n=5 Control and n=3 Family RoMS lines. Data are expressed as mean ± SEM, where individual dots represent the mean for each cell line. (g) Quantification of iPericyte cell area. > 50 cells were measured per line per treatment condition for n=5 Control and n=4 Family RoMS lines. Data are expressed as median ± interquartile range, and individual dots represent the cell line means. (h) Quantification of the mean velocity of iPericytes under untreated and myelin conditions. Velocity was determined for > 10 cells per line per treatment. Data are expressed as median ± interquartile range, and individual dots represent cell line means. (i) Quantification of nucleus displacement between t=0 and t=60 minutes of imaging for iPericytes under untreated or myelin conditions. Displacement was measured for > 10 cells per line per treatment for n=4 Control and n=4 Family RoMS lines. Data are expressed as median ± interquartile range, and individual dot points represent cell line means. (j) Principal component analysis of gene expression in untreated or myelin treated iPericytes. (k) Volcano plot comparing gene expression in iPericytes cultured without or with myelin debris. Adjusted *p*-value <0.05 and Log2 fold change ≥ 0.585; n=9 (n=5 Control and n=4 Family RoMS) lines. Restricted maximum likelihood linear mixed model with pairwise comparisons using Tukey’s adjustment for p-values. False discovery rate adjustment was used for multiple testing for RNAseq data. _∗∗∗∗_p<0.0001.

To determine whether 44 hours of exposure to myelin debris results in transcriptional changes in Control and Family RoMS iPericytes, we performed bulk RNA sequencing. PCA of global gene expression indicated that cell line was the primary source of variation, and that exposure to myelin debris did not have a significant effect on gene expression (**Fig. 3j**). Differential gene expression analysis confirmed that myelin phagocytosis did not change the iPericyte transcriptional profile, and no genes showed evidence of an interaction effect between myelin treatment and disease status (**Fig. 3k; Fig. S4a-b**; **Data TS8-10**). These data indicate that pericytes respond differently than astrocytes, microglia, and brain microvascular endothelial cells, each of which change their proliferation rate and gene expression profile with myelin debris exposure ^56,58,59^. However, our data do not rule out the possibility that the duration of myelin exposure, whether acute or sustained, could alter the pericytes, or that myelin exposure alters protein translation or induces post-translational modifications in pericytes rather than regulating gene transcription.

### Hypoxia increases iPericyte proliferation, morphological remodelling, and angiogenic signalling

The MS lesion environment can become hypoxic, and this is likely to induce hypoxic injury ^14,15,17,60,61^. Pericytes are early responders to brain hypoxia, commencing migration along vessels within 2 hours of a hypoxic injury ^62^. To determine how hypoxia impacts iPericyte behaviour and determine whether MS status alters the hypoxic response, we cultured Control and Family RoMS iPericytes under normoxic or hypoxic (0.5% O_2_) conditions for 20 hours. Normoxic and hypoxic iPericytes incorporated EdU (**Fig. 4a**), and the proportion of iPericytes that were EdU^+^ increased significantly under hypoxic conditions (**Fig. 4b**). This response is consistent with that of immortalised retinal pericytes, which also increase their proliferation rate when cultured with <1% O_2_ ^63^. However, the hypoxia-induced increase in proliferation was equivalent for Control and Family RoMS iPericytes (**Fig. 4b**), indicating that this response is unaltered by RoMS status.

**Figure 4:**
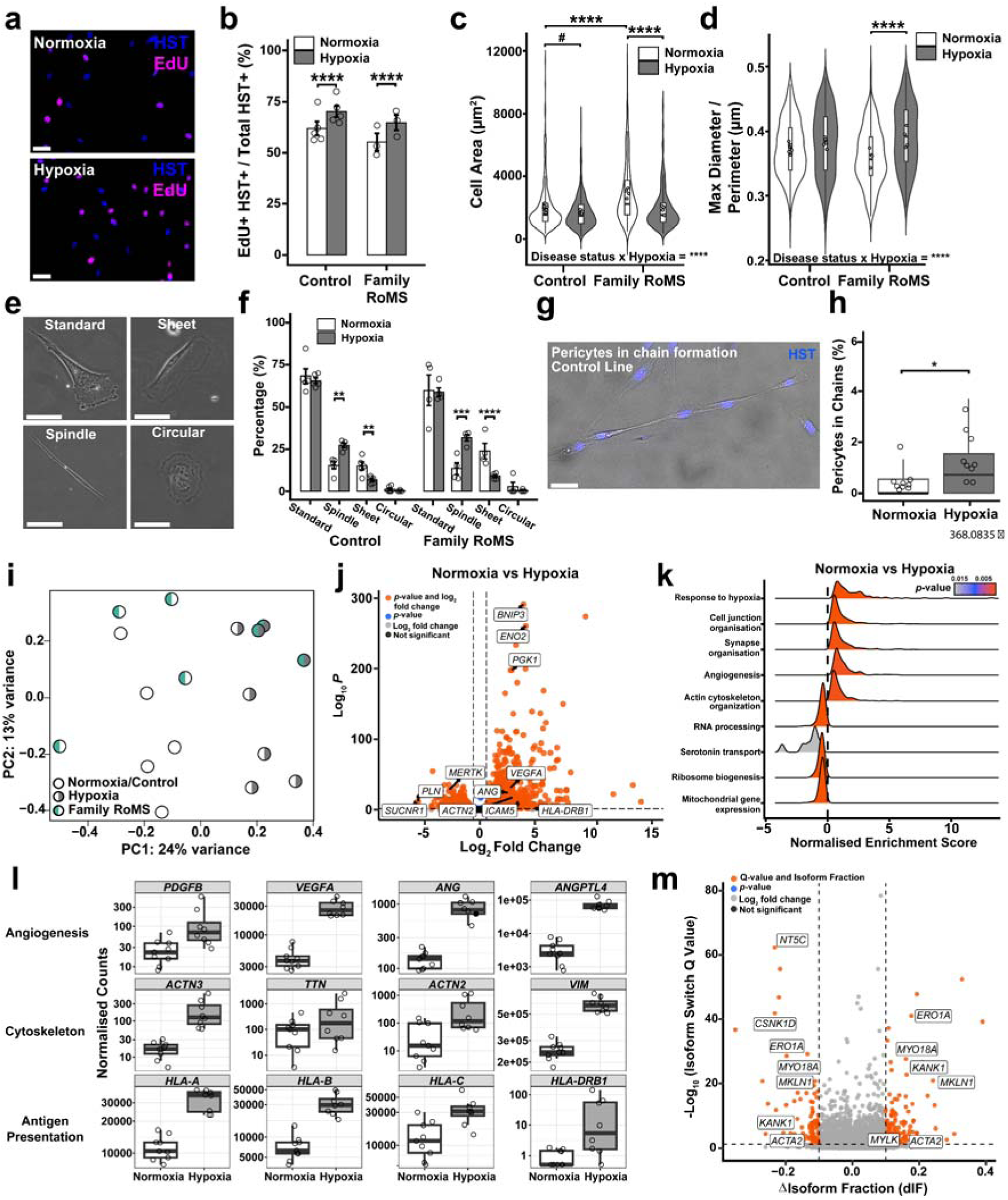
Hypoxia induces proliferation and alters the morphology and transcriptomic profile of iPericytes. (a) Images (10x) of Control iPericytes that incorporate EdU (magenta) when cultured under normoxic or hypoxic (0.5% O_2_) conditions for 20 hours. Scale bars, 50µm. (b) Quantification of the percentage of cells in Control or Family RoMS iPericyte cultures that were EdU^+^ after 20 hours of exposure to a normoxic or hypoxic environment. 4-9 wells analysed per line per treatment for n=5 Control and n=3 Family RoMS lines. Data are expressed as mean ± SEM, and individual dots represent cell line means. (c,d) Quantification of iPericyte cell area or max Feret diameter (max diameter/perimeter) for Control and Family RoMS iPericytes grown under normoxic or hypoxic conditions. > 50 cells measured per cell line per treatment for n=10 Control and n=4 Family RoMS lines. Data are expressed as median ± interquartile range, and individual dot points represent cell line means. (e) Images (20x) showing example Control iPericyte morphologies: standard, spindle, sheet and circular. Scale bars, 50 µm. (f) Quantification of the percentage (%) of iPSC pericytes in Control or Family RoMS cultures that adopted each morphological subtype when cultured under normoxic or hypoxic conditions. > 22 cells were measured per line per treatment for n=5 Control and n=4 Family RoMS lines. Data are expressed as mean ± SEM, individual dot points represent cell line means and was analysed using a binomial generalised linear mixed model. (g) Image (10x) of Hoechst^+^ Control iPericytes cultured under hypoxic conditions, forming a chain. Scale bar, 50µm. (h) Quantification of the percentage (%) of iPericytes forming chains under normoxic and hypoxic conditions. 4-9 wells analysed per line per treatment for n=9 (n=5 Control, n=4 Family RoMS) lines. Data for Control and Family RoMS lines were combined as the response was equivalent. Data are expressed as median ± interquartile range, and individual dot points represent cell line means. (i) Principal component analysis of gene expression in iPericytes cultured under normoxic or hypoxic conditions (n=5 Control, n=3-4 Family RoMS cell lines per treatment condition). (j) Volcano plot showing genes that are differentially expressed between iPericytes cultured under normoxic or hypoxic conditions. Adjusted p-value <0.05 and Log2 fold change ≥0.585; data from n=8 (n=5 Control and n=3-4 Family RoMS) lines were combined as MS status did not impact response to hypoxia. (k) Ridge plot showing gene set enrichment analysis results comparing normoxic and hypoxic iPericytes. (l) Boxplots showing gene expression in normoxic and hypoxic iPSC pericytes, for differentially expressed genes associated with angiogenesis, cytoskeletal regulation and antigen presentation. Data are expressed as median ± interquartile range, and individual dot points represent cell line means (n=5 Control and n=3-4 Family RoMS per treatment condition). (m) Volcano plot showing genes that have isoform switches between iPericytes cultured under normoxic or hypoxic conditions (n=5 Control and n=3-4 Family RoMS per treatment condition). Restricted maximum likelihood linear mixed model with pairwise comparisons using Tukey’s adjustment for p-values. #<0.1, _∗_p<0.05, _∗∗_p<0.01, _∗∗∗∗_p<0.0001. False discovery rate adjustment was used for multiple testing for RNAseq data.

Control and Family RoMS cells were smaller under hypoxic conditions (**Fig. 4c**), which is consistent with previous *in vitro* and *in vivo* reports that hypoxia induces pericyte contraction ^25,64,65^. However, we also determined that more of the Control and Family RoMS iPericytes adopted a spindle morphology under hypoxic conditions (**Fig. 4d-f**) and organised themselves into chains of 3-8 cells (**Fig. 4g**). Only ∼0.5-3.5% of individual iPericytes formed chains in Control or Family RoMS cultures, yet this produced a striking change in the culture appearance (**Fig. 4h**). The iPericyte chains recapitulate the end-to-end organisation of pericytes along blood vessels *in vivo* ^66^ and likely reflect their pro-angiogenic role in providing scaffolding for new vessels and supporting basement membrane extension. ^32^. Consistent with this idea, hypoxic Control and Family RoMS iPericytes were more motile, increasing their velocity and nuclear displacement (**Fig. S5a,b**), as previously reported for retinal pericytes ^63^. These hypoxia-driven changes in pericyte motility and organisation may help explain the hypervascularisation observed in MS ^67,68^.

To uncover transcriptional changes that underpin the hypoxia-induced morphological and behavioural changes, we performed bulk RNA sequencing on Control and Family RoMS iPericytes cultured under normoxic and hypoxic conditions. Hypoxia accounted for ∼24% of variation (PC1), and samples did not separate by MS status (**Fig. 4i**; **Fig. S4c, Data TS11**). Our differential gene expression analysis, comparing Control and Family RoMS iPericytes under hypoxic conditions revealed the same DEGs identified in our baseline comparison (**Fig. S4d, Data TS12**). By comparing iPericytes cultured under normoxic or hypoxic conditions, independent of MS status, we identified >1,900 DEGs (log fold change threshold >0.585), and most were upregulated with hypoxia (**Fig. 4j, Data TS13**). Of note, the transcriptional response to hypoxia was not modified by exposure to myelin debris (**Fig. S4e**). To determine whether the differential gene expression was being driven by particular transcription factors that were activated under hypoxic conditions, we used decoupleR to calculate transcription factor enrichment scores. We determined that many of the transcriptional changes induced by hypoxia, resulted from increased hypoxic inducible factor (HIF)1α activity (**Fig. S5c-f**) and reduced sterol regulatory element binding (SREB) protein activity (**Fig. S5c**). As the degradation of SREB transcription factors reduces the oxygen-intensive process of cholesterol synthesis ^69^, this likely represents a pro-survival response.

Gene set enrichment analysis, using gene ontology (GO) biological processes, identified 176 significantly enriched terms (*p-adjusted*<0.05; **Data TS14**). Consistent with the morphological and behavioural changes observed in hypoxic iPericytes, enriched GO terms included “response to hypoxia”, “angiogenesis” and “actin-cytoskeleton organisation” (**Fig. 4k**). Notably, *ANGPTL4* was upregulated (**Fig. 4l**) and was annotated to both the hypoxia and angiogenesis pathways. *ANGPTL4* encodes angiopoietin-like 4, a secreted factor that regulates neovascularisation, pericyte coverage, and BBB integrity ^70,71^. The increased expression of *ANGTPL4* and other angiogenic genes, such as *ANG* and *VEGFA*, could account for the elevated iPericyte motility and altered morphology (**Fig. 4c,d,f; Fig. S5a,b**). Furthermore, the upregulation of cell cycle and proliferation genes, including *PDGFB* and *CDK6*, (**Fig. 4l, Fig. S5g**), may explain the small but significant increase in iPericyte proliferation with hypoxia (**Fig. 4b**). A marked increase in antigen presentation genes, including MHC class I (*HLA-A, -B, -C*) (**Fig. 4l**) suggests that iPericytes are signalling increased cellular stress or injury, and increasing their visibility to the immune system ^72^. By contrast, induction of MHC class II (*HLA-DRB1*) (**Fig. 4l**) reflects a more profound shift of the cells to an acquired, non-canonical antigen-presenting state, typically restricted to professional immune cells ^73,74^. These changes indicate that pericytes in hypoxic MS lesions may adopt an immunomodulatory phenotype capable of shaping local inflammatory activity and influencing leukocyte interactions with the neurovasculature ^75,76^.

Hypoxia suppressed the transcription of genes related to mitochondria and ribosome function, and RNA processing (**Fig. 4k**). It also altered transcript usage for 185 genes (**Fig. 4m, Data TS15**) - most of which were not differentially expressed genes (**Fig. S5h**). As an example, hypoxia did not alter overall *MYLK* expression, but was associated with a reduction in the short Telokin-encoding transcript (padj=0.0035) (**Fig. S5i**). This would be predicted to promote contraction and may partly explain the reduction in iPericyte size.

### Cytokines increase ET-1-evoked iPericyte contractility

Within MS lesions, pericytes would be exposed to pro-inflammatory cytokines, such as TNFα and IFNγ ^77,78^. To determine how this could modify iPericytes behaviour, we cultured Control and Family RoMS iPericytes with 20ng/mL TNFα, 40ng/mL IFNγ, or both for 20 hours. The combined TNFα+IFNγ treatment, but not either cytokine alone, reduced iPericyte proliferation, irrespective of MS status (**Fig. 5a,b**). Exposure to TNFα+IFNγ also increased the cell area and perimeter of Control iPericytes (**Fig. 5c, Fig. S6a**) such that their size resembled that of untreated Family RoMS iPericytes. The failure of Family RoMS iPericytes to change in size with TNFα+IFNγ treatment may reflect a ceiling effect, as they started from a larger baseline size, or an impaired response to cytokine signalling. However, cytokine treatment did not produce a change in overall shape, as maximum Feret diameter was unchanged (**Fig. S6b**).

**Figure 5:**
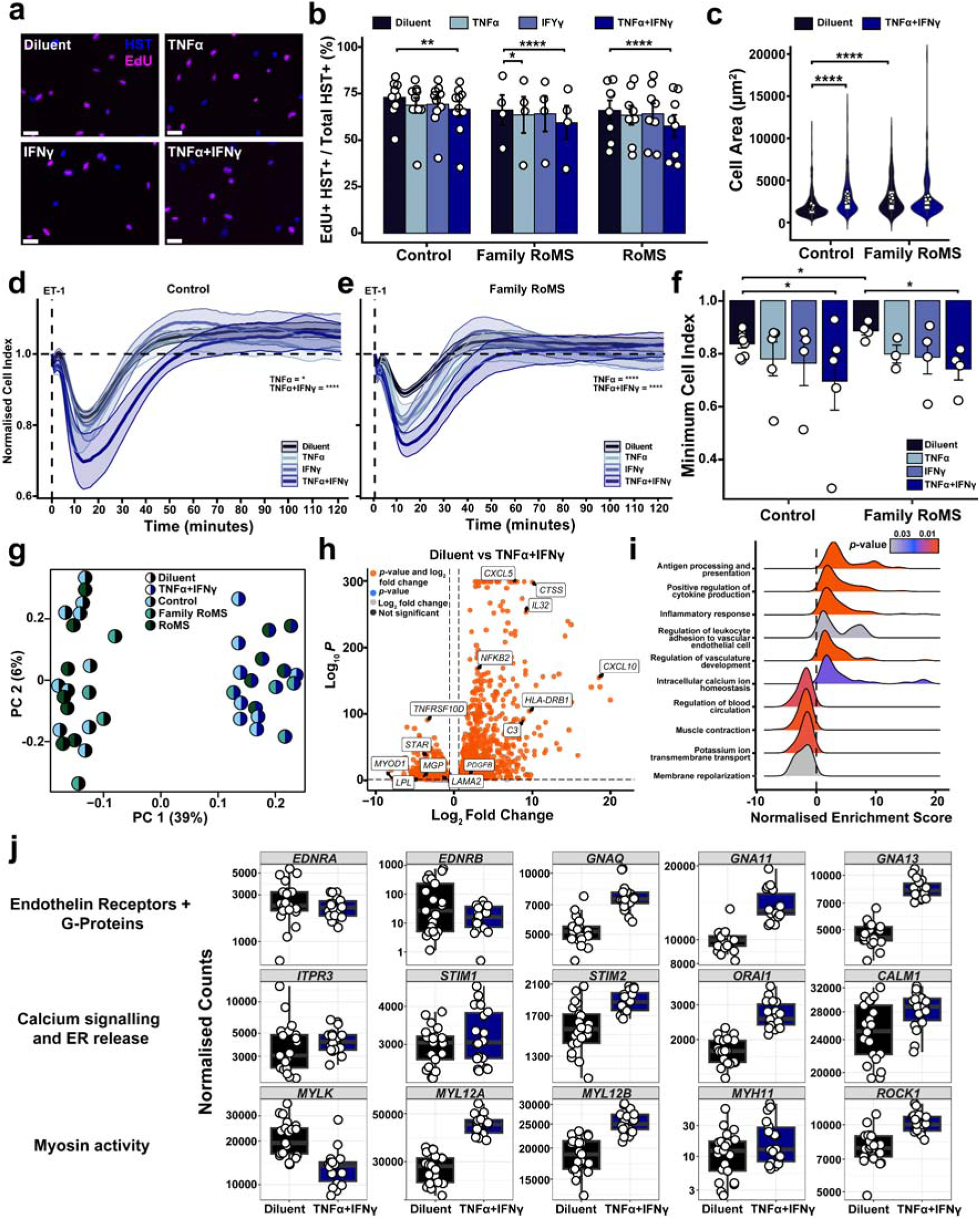
MS-associated inflammatory cytokines reduce proliferation, increase cell size and enhance contractility of iPericytes. (a) Images (10x) of EdU^+^ (magenta) Control iPericytes following 20 hours of diluent, TNFα, IFNγ or TNFα+IFNγ treatment. Scale bars, 50µm. (b) Quantification of the percentage (%) of cells within Control, Family RoMS and RoMS iPericyte cultures that incorporated EdU when cultured with diluent, TNFα, IFNγ or TNFα+IFNγ for 20 hours. 4-9 wells were analysed per line per treatment for n=10 Control, n=4 Family RoMS and n=8 RoMS cell lines. Data are expressed as mean ± SEM, and individual dot points represent cell line means. (c) Quantification of Control, Family RoMS and RoMS iPericyte area for cells cultured with diluent or TNFα+IFNγ. >50 cells were measured per line per treatment for n=10 Control and n=4 Family RoMS lines. Data are expressed as median ± interquartile range, and individual dot points represent line mean. (d,e) Normalised electrical impedance cell index of Control and Family RoMS iPericytes cultured with diluent, TNFα, IFNγ or TNFα+IFNγ for 20 hours prior to exposure to ET-1. Data are normalised to cell index at the time of endothelin-1 addition. 2-7 wells were analysed per cell line for n=5-10 Control and n=3-4 Family RoMS lines. Data are expressed as mean ± SEM. (f) The normalised minimum cell index (maximum contraction) after treating Control or Family RoMS iPericytes with diluent, TNFα, IFNγ or TNFα+IFNγ. 2-7 wells were evaluated per cell line for n=5-10 Control and n=3-4 Family RoMS lines. (g) Principal component analysis of gene expression in iPericytes cultured with diluent or TNFα+IFNγ treatment (n=8-10 Control, n=4 Family RoMS and n=6-7 RoMS cell lines per treatment condition). (h) Volcano plot showing genes that are differentially expressed between iPericytes cultured with diluent or TNFα+IFNγ treatment. Adjusted p-value <0.05 and Log2 fold change ≥0.585; data from n=18 (n=8 Control and n=4 Family RoMS and n=6 RoMS) lines were combined as MS status did not impact response to TNFα+IFNγ. (i) Ridge plot showing gene set enrichment analysis results comparing diluent and TNFα+IFNγ treated iPericytes. (j) Boxplots showing gene expression from diluent and TNFα+IFNγ treated iPericytes, for differentially expressed genes associated with endothelin receptors, calcium signalling and myosin activity. Data are expressed as median ± interquartile range, and individual dot points represent cell line means (n=8-10 Control, n=4 Family RoMS and n=6-7 RoMS cell lines per treatment condition). Data are expressed as mean ± SEM. Restricted maximum likelihood linear mixed model with pairwise comparisons using Tukey’s adjustment for p-values. _∗_p<0.05, _∗∗_p<0.01, _∗∗∗∗_p<0.0001. False discovery rate adjustment was used for multiple testing for RNAseq data

To determine whether inflammatory cytokines influence iPericyte contractility and, by extension, vascular tone, we initially exposed a Control iPericyte line to TNFα+IFNγ for 0.5-20 hours, before assessing the response to ET-1. iPericyte contraction was enhanced after 1 hour of TNFα+IFNγ exposure but was increased ∼2-fold by 20 hours (**Fig. S6c**). We therefore exposed Control and Family RoMS iPericytes to TNFα, IFNγ or both for 20 hours, before measuring the ET-1-evoked contraction. As expected, the Control iPericyte contraction was larger than the Family RoMS iPericyte contraction at baseline (**Fig. 5d,e,f**). Exposure to TNFα or IFNγ alone increased the maximum Control or Family RoMS ET-1-evoked contraction by 44.2% and 86.9% (TNFα) or 34.4% and 76.9% (IFNγ), respectively, although IFNγ did not reach statistical significance (**Fig. 5d,e,f**). The combined TNFα+IFNγ treatment had an additive effect, increasing the maximum contraction of Control and Family RoMS iPericytes by 85.5% and 125.9%, respectively (**Fig. 5d,e,f**), such that the maximum contraction size became equivalent for Control and Family RoMS iPericytes (**Fig. 5f**). The TNFα+IFNγ stimulated iPericytes also took longer to return to their baseline size (**Fig. 5d,e**). A larger and sustained pericyte contraction would greatly restrict blood flow within active and smouldering lesions which would, in turn, exacerbate cellular metabolic stress, promote neurodegeneration, and impede repair ^79,80^ (**Fig. 6**).

**Figure 6:**
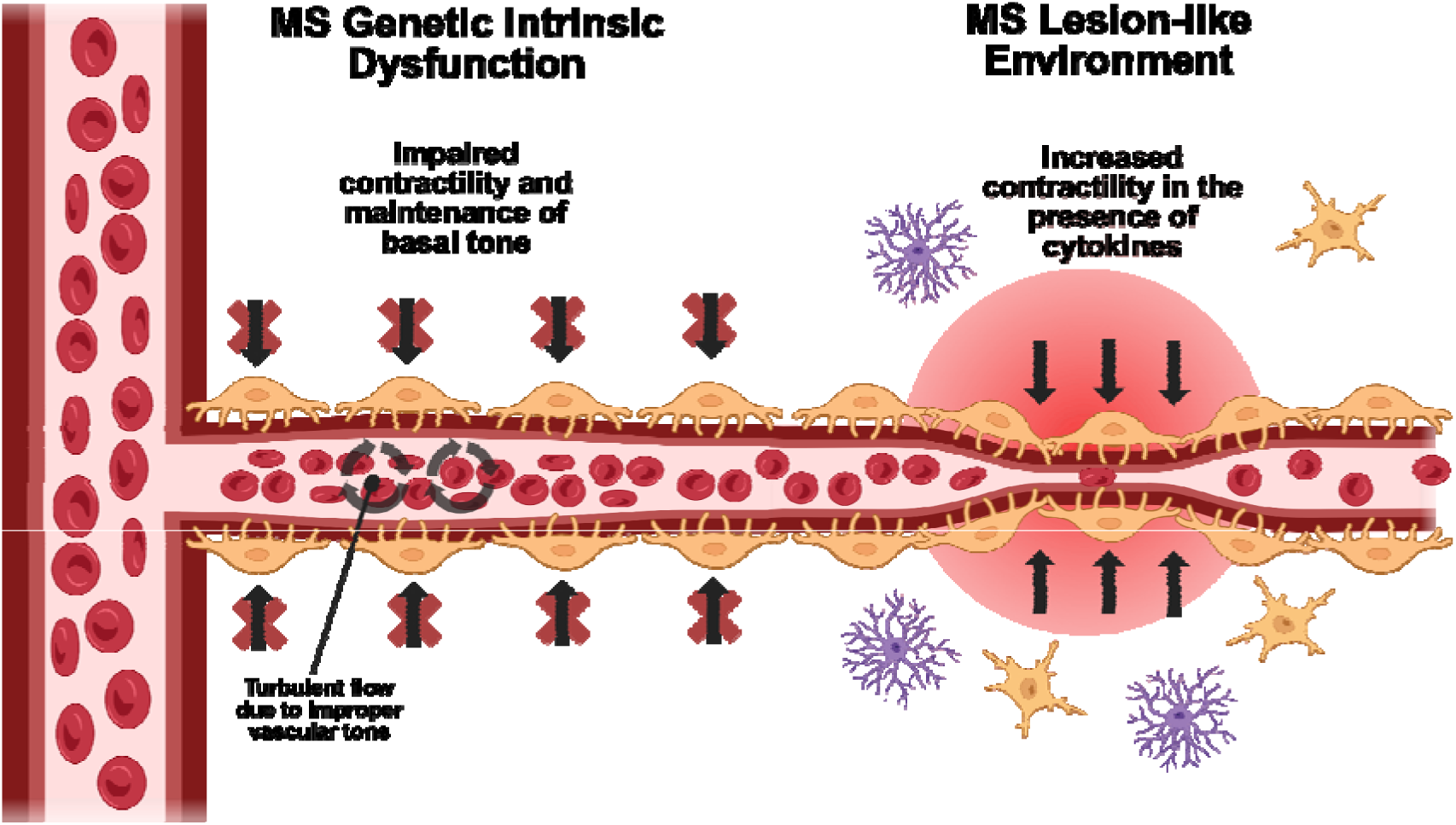
Pericyte dysfunction in multiple sclerosis. Schematic illustrating how intrinsic pericyte dysfunction and heightened contractile responses to an MS lesion–like inflammatory environment converge to disrupt vascular tone and impair cerebral capillary blood flow.

To determine how TNFα+IFNγ could exert these effects, we performed bulk RNA sequencing. PCA revealed that TNFα+IFNγ exposure accounted for ∼39% of gene expression variation (PC1), and the samples did not cluster based on MS status (**Fig. 5g**). Consistent with these data, we did not detect an interaction between MS status and cytokine exposure for any gene (**Fig. S6d, Data TS16**) and differential gene expression analysis comparing treated Control and either Family RoMS or RoMS iPericytes, only identified differences previously described for baseline conditions (**Fig. S6e,f, Data TS17,18**). By contrast, TNFα+IFNγ exposure altered the expression of >3,800 genes (**Fig. 5h, Data TS19**). Transcription factor analysis revealed that NFKB1 and IRF1 signalling were activated (**Fig. S6g,h**). NFKB1 and IRF1 activation reflects the canonical responses to TNFα and IFNγ, results in the increased expression of many chemokine and complement genes and is likely to result in pericytes promoting leukocyte extravasation in the CNS (**Fig. S6i**). By contrast, PPARG and MYOG transcriptional networks were downregulated (**Fig. S6g,h**), indicating that lipid metabolism and anti-inflammatory signalling are suppressed, along with cytoskeletal maintenance. Gene set enrichment revealed that cytokine treatment resulted in iPericytes activating angiogenesis and antigen presentation pathways, but downregulating blood circulation and calcium signalling pathways (**Fig 5i, Data TS20**). Our analyses of the transcriptomics changes induced by treatment with TNFα or IFNγ alone (**Fig. S7a,b, Data TS22,23**) indicate that TNFα signally largely accounts for the upregulation of genes involved in kinase activity and leukocyte migration, while IFNγ modifies cytokine production and alters the innate immune response (**Fig.S7a-f, Data TS24,25,26**).

To determine how cytokine treatment modified the ET-1 signalling pathway (**Fig. S3b**) to enhance iPericyte contraction, we examined DEGs within the ‘regulation of blood circulation’ pathway. Notably, expression of the ET-1 receptors, *EDNRA* and *EDNRB*, remained unchanged (**Fig. 5j**), suggesting that the enhanced contractile response resulted from modifications in downstream signalling. Consistent with this, the G-protein alpha subunits that are coupled to ET-1 receptors (*GNAQ*, *GNA11*, and *GNA13,* **Fig. 5j**) ^81^ were upregulated, along with genes that influence calcium dynamics and endoplasmic reticulum calcium release, and are critical for an evoked and sustained contraction (*ITPR3*, *STIM1*, *STIM2*, *ORAI1*, and *CALM1*, **Fig. 5j**). In parallel, key components of the downstream actomyosin contractile apparatus were elevated (*MYL12A*, *MYL12B*, *MYH11* and *ROCK1*, **Fig. S3b**). Our DTU analysis also identified 517 genes (**Fig. S6j, Data TS21**). Of note, total *MYLK* gene expression was decreased (**Fig. 5j**) due to selective downregulation of the telokin transcript (**Fig. S6k**). Reduced telokin expression would be predicted to increase MLCK activity and myosin phosphorylation to enhance contractility. These transcriptional changes indicate that cytokines modify multiple downstream mediators of pericyte contraction, to drive an enhanced and sustained contractile response that would restrict capillary blood flow within MS lesions.

## Discussion

Herein, we show that RoMS iPericyte experience intrinsic changes in gene expression that impacts key contractile mediators, including *MYLK* (**Fig. 1**). Consistent with this, RoMS iPericytes are larger, and have a reduced contractile response to the vasoconstrictor, ET-1 (**Fig. 2**). While intrinsic dysfunction has the potential to initiate or exacerbate MS pathogenesis, Control and Family RoMS iPericytes responded similarly to MS-relevant environmental stimuli. Control and Family RoMS iPericytes phagocytosed myelin debris (**Fig. 3**) and responded to hypoxia by proliferating and upregulating transcripts that promote angiogenesis and contractility (**Fig. 4**). Exposure to the cytokines TNFα and IFNγ resulted in larger and longer ET-1 evoked contractions, that were associated with transcriptional changes expected to enhance ET-1 receptor G-protein coupled signalling, intracellular calcium regulation, the actomyosin contractile machinery, and MLCK activity (**Fig. 5**). These data indicate that prior to MS onset, pericyte dysfunction would result in capillary dilation across the CNS and likely hypoperfusion, while disease onset and lesion formation would drive severe pericyte contraction and capillary constriction (**Fig. 6**).

Hypoperfusion and impaired neurovascular coupling may precede MS clinical onset and even contribute to lesion development ^27^. Individuals with clinically isolated syndrome have an increased cerebral blood volume that has a longer mean transit time, corresponding to reduced cerebral perfusion ^13,55^. Our data suggest that this is, at least in part, due to an impaired ability of pericytes to contract appropriately in response to vasoactive signals. Local vasodilation is typically associated with an increase in regional blood flow. However, when this is not successfully paired with the vasoconstriction of capillary beds elsewhere in the cerebrovascular network, it could instead result in hypoperfusion and could fail to direct blood to the most active brain regions ^26,82^. This is demonstrated experimentally by pericyte depletion studies, which show that the loss of pericytes from across the capillary bed, results in increased capillary blood volume, but global cerebral hypoperfusion ^36,83^. Indeed, the focal ablation of 3 proximal pericytes is sufficient to stall blood flow and cause capillary regression ^82^. Disturbances in blood flow can also alter endothelial cell phenotypes, causing endothelial cells to upregulate genes involved in immune cell chemotaxis and inflammation ^84^. As iPSC-derived endothelial cells from people with MS express elevated levels of the leukocyte adhesion molecules, ICAM-1 and VCAM-1 ^24^, pericyte dysfunction that induces blood flow stalling would further exacerbate immune cell adhesion to and migration across the BBB. Importantly, adhesion of leukocytes to brain capillaries would further reduce blood flow ^85^. Consequently, the disrupted blood flow from impaired pericyte-mediated vasoconstriction and intrinsic endothelial cell alterations may create a vascular environment that is more permissive to immune cell infiltration, directly promoting MS development.

Once MS develops, it is likely that the intrinsic pericyte dysfunction becomes overshadowed or masked by maladaptive responses to pathophysiological processes such as hypoxia and inflammation. Although people with MS exhibit a typical cerebral blood volume ^55^, they show reduced cerebral blood flow ^7,11,12^. This reduction in blood flow is often accompanied by hypervascularisation ^67,68^, which likely arises as a secondary compensatory response to reduced blood flow and the resulting hypoxia ^14,15^. Our data demonstrate that exposure to 0.5% O_2_ induces pericyte proliferation and morphological changes that are accompanied by the upregulation of cell cycle, cytoskeletal, and angiogenic genes, and this response could readily contribute to the hypervascularisation observed in people with MS. In particular, the increase in spindle shaped pericytes and the formation of interconnected chains likely reflect a shift toward a pro-angiogenic state. We hypothesise that this is a monoculture *in vitro* presentation of the scaffolding role pericytes have to support endothelial tip cells, and direct new blood vessel formation *in vivo* ^32,33,86^. Despite higher vessel densities in the MS brain ^67,68^, hypoperfusion is still observed, likely due to chronic inflammation resulting in poor vessel development, hyper-constriction via pericytes and damage.

In MS lesions, the elevated levels of TNFα ^78^ and IFNγ ^77^ would significant impair pericyte function. When iPericytes were exposed to TNFα ^78^ and IFNγ that upregulated genes involved in G-protein, calcium and actomyosin signalling, and experienced larger and longer ET-1 evoked contractions. These cytokine-induced changes to pericyte function could readily explain the narrowed capillaries, reduced blood flow and heterogenous capillary transit times reported within MS lesions ^18,87,88^. However, the ability of cytokines to increase the size and duration of pericyte contraction also has implications for neurovascular dysfunction in other neuroinflammatory conditions, including stroke, where it likely contributes to the no-reflow phenomenon ^65,89^. As cytokine-stimulated iPericytes also increase their expression of chemokine genes, they may actively promote leukocyte recruitment to the CNS, which could promote capillary plugging and exacerbate hypoperfusion ^85^.

In people with MS, cerebral blood flow is inversely associated with T1 lesion volume, neuroinflammation, and serum neurofilament light chain (neurodegeneration) ^8,79,80^, indicating that blood flow restoration could have a significant impact on neuroprotection and repair. The potential benefit of restoring perfusion is highlighted by studies in EAE and Alzheimer’s disease (APP^NL–G–F^) mice, where treatment with the CNS-selective vasodilator nimodipine improved perfusion and reduced demyelination (EAE only), hypoxia, immune cell stalling, and neurological deficits ^90,91^. These findings underscore the importance of targeting pericyte function and cerebral perfusion as a therapeutic strategy in MS.

## Methods

### Resource availability

Further information and requests for data or resources related to this publication should be directed to the corresponding author.

#### iPSC line generation and availability

All iPSC lines used in this study were generated for our MS Stem biobank (Menzies Institute for Medical Research) and can be purchased from https://msresearchflagship.org.au/researchers/ms-stem. The iPSC lines were generated and characterised as previously described ^40,41^. The generation and use of each iPSC line was approved by the University of Tasmania Human Research Ethics Committee (approvals 16915 and 30092). All study participants provided informed consent for the blood sample collection, iPSC generation, differentiation, and genetic data sharing conducted as part of this project. n=10 Control, n=4 Family ^40^ and n=8 RoMS iPSC lines (**Table 1**) were used in this study.

#### iPSC culture and iPericyte differentiation

iPSCs were cultured on Matrigel^®^ (Corning) coated 60mm Nunclon Delta plates (ThermoFisher) in mTeSR1™ Plus medium (StemCell Technologies, cat.#100-0276). iPSCs were passaged every 6-8 days and the medium exchanged every 2 days. One day prior to passaging, areas of spontaneous differentiation were removed by scraping the plate with a 200µl pipette tip and washing with DMEM/F12 (ThermoFisher). For passaging, cells were dissociated into clumps by incubating cells in ReLeSR^TM^ (StemCell Technologies, cat.#100-0484). iPericytes were generated as previously described ^39^, by plating iPSCs in 60mm matrigel-coated plates. After 2 days, the medium was exchanged for STEMdiff^TM^ Mesoderm Induction Medium (MIM; StemCell Technologies, #05221), replaced daily for 5 days, and complete pericyte medium (CPM) containing pericyte growth supplement and fetal bovine serum (FBS; ScienCell, #1201), replaced daily for the subsequent 5 days. On day 10, the cells were dissociated with Accutase^®^ (StemCell Technologies, #07922) and plated in uncoated 6-well plates at a density of 5 x 10^4^ per cm^2^. The medium was replaced every 48h, and the cells passaged at ∼80% confluency (∼5-7 days). iPericytes were frozen in CPM with 10% dimethyl sulfoxide and stored in liquid nitrogen. iPericytes were thawed into CPM and culture for 2 weeks prior to use in experiments. Passage 3-7 iPericytes were used for experiments.

#### Immunocytochemistry

Pericytes were grown to ∼25% confluency in 24 well plates and fixed with cold 4% (w/v) paraformaldehyde (Sigma) in PBS for 15 mins at 21°C. Cells were washed with PBS and then incubated in blocking solution (10% Tween-20 / 2% FBS in PBS) for 30 minutes at 21°C. Primary antibodies (goat anti-PDGFRβ, R&D Systems AF385, RRID:AB_777165; rabbit anti-CD13, Abcam Ab108310, RRID:AB_10866195;) were diluted in the blocking solution and applied overnight at 4°C, and the plates placed on an orbital shaker. Cells were washed thrice in PBS before secondary antibodies (Alexa Fluor 488-conjugated donkey anti-rabbit; Alexa Fluor 647-conjugated donkey anti-goat) and Hoechst 33342, diluted in blocking solution, were applied for 2 hours at 21°C. Cells were washed thrice in PBS and maintained in PBS for imaging.

#### Fluorescent Microscopy

All fluorescent images were acquired using an iXon Ultra 888 EMCCD camera mounted on an inverted Nikon Eclipse Ti microscope, with imaging performed blinded to disease status and/or treatment group. Either a Plan Apo 10× or Plan Apo VC 20× DIC N2 objective was used. Illumination was provided by a SPECTRA X light source equipped with Semrock 387/11, 485/20, 560/25, and 650/13 excitation filters, a Semrock FF410/504/582/669 dichroic mirror, and Semrock 440/521/607/700 emission filters. Uniform exposure times were maintained across all images within each experiment.

#### RNA sequencing

iPSCs were seeded on 60mm plates and cultured to ∼75% confluency prior to collection. Pericytes were seeded on 6-well plates (300,000 cells per well) and cultured for 44 hours prior to collection. Pericytes were exposed to hypoxic conditions or cytokine treatment for 20 hours. They were exposed to myelin debris for 44 hours. Cells were dissociated using Accutase, washed in cold DPBS and lysed using RLT buffer (Qiagen). The lysate was stored at −20°C and thawed on ice for RNA extraction using the RNeasy Mini Kit, following the manufacturer’s instructions (Qiagen, #74106). Briefly, samples were transferred to RNeasy spin columns, DNA removed by treatment with DNase (Qiagen, #79254) and RNA eluted in 40µl of RNase free water. RNA concentration and integrity were evaluated using the Agilent 4200 Tapestation system. Briefly, 5µl of RNA ScreenTape Sample Buffer (Agilent, #5067-5577) and 1µl of RNA sample or ladder were combined by vortexing at 2,000rpm for 1 minute using an IKA vortex. Samples were incubated at 72°C for 3 minutes, placed on ice for 2 minutes, loaded into the TapeStation and run on RNA ScreenTape (Agilent, #5067-5576) via electrophoresis. Samples with > 20ng/µL RNA and a RNA integrity number (RIN) > 8 were sent to the Australian Genome Research Facility for bulk RNA sequencing to a minimum depth of 20 or 50 million reads with 150 base paired end lengths. Libraries were generated with the Illumina Stranded mRNA workflow with polyA capture. Raw RNA sequencing data is available for download from the European Genome Phenome Archive (accession to be provided for publication) or, for data generated from a limited number of the iPSC lines, by reasonable request to the corresponding author.

#### Re-analysis of publicly available RNAseq datasets

HBVP, iPSC and iPericyte raw bulk RNA sequencing data from King, et al. ^39^ were downloaded from GEO (accession GSE252046) and the European Genome Phenome Archive (accession EGAD50000000255).

#### RNA data processing

Raw sequencing data were processed using nf-core/rnaseq v3.12.0 ^92^, and a nf-core workflow executed with Nextflow v.23.04.2. Initially, the sequencing adapter (5’-CTGTCTCTTATACACATCT - 3’) was trimmed from each read and 1 base pair removed from the 5’ end of each read using TrimGalore ^93^. A read quality Phred score threshold of 20 was used to remove low quality reads. Reads <25 base pairs after trimming were discarded. The quality of the sequencing data was evaluated using FastQC (v.0.11.9), and all data met the necessary quality metrics for analysis. Gene expression was quantified using Salmon in mapping-based mode (v.1.10.1) with the *Homo sapiens* (Hg38) reference transcriptome. The sequencing library type was inward stranded reverse (ISR). Within Salmon, --seqBias and --gcBias were employed to enable Salmon to learn and correct for sequence-specific biases and fragment-level GC biases, respectively. The result of this analysis was per sample quantification of transcript expression.

#### Differential gene expression analysis, differential transcript usage and gene set enrichment

Differential gene expression analysis was performed using DESeq2 ^94^ (v1.42.0). Tximport ^95^ (v.1.30.0) was used to import transcript-level abundance into R (v.4.4.2) and summarise transcript abundance to the gene-level. Genes with <10-100 reads across each group were filtered out. A first pass differential gene expression was performed comparing gene expression levels between the experimental groups being tested. Factors of unwanted variation were estimated and included as covariates in the model using RUVSeq ^96^ (v1.36.0) as required. A *p*-value of 0.5 was used to define a set of genes not differentially expressed in the first pass analysis for use as in-silico empirical negative control genes in RUVSeq. Differential gene expression was tested under a hypothesis of log_2_ fold changes > 0.1 for Control vs RoMS testing and 0.585 (corresponding to a 1.5-fold change in expression) for treatment testing, with a false discovery rate threshold of alpha <0.05. When examining interaction effects between disease status and treatment a log_2_ fold change threshold of > 0.1 was used. When examining interaction effects between TNFα and IFNγ treatment a log_2_ fold change threshold of > 0.585 was used. Each differential gene expression analysis was visualised using EnhancedVolcano ^97^ (v.1.20.0), pheatmap ^98^ (v.1.012) or ggplot2 ^99^ (v.3.5.1). Differential transcript usage analysis was performed using IsoformSwitchAnalyzer (v.2.6.0) ^100^ with a gene expression cut off of 10 and isoform expression cut off of 3. DEXSeq (v.1.52.0) ^101^ was used for DTU statistical testing. Gene set enrichment analysis (GSEA) was completed using clusterProfiler (v.4.14.4) ^102^. Transcription factor analyses were completed using decoupleR ^103^ (v.2.8.0).

#### EdU proliferation assay

EdU assays were performed as previously described ^39,104^, between passage 4 and 5. Briefly, pericytes were plated into 96-well plates at 5,000 cells per well and left to adhere overnight. The following day, the medium was replaced with CPM containing 1µM EdU (ThermoFisher) and the cells incubated for 20 hours. The medium was then removed and the cells fixed with cold 4% paraformaldehyde in PBS for 15 minutes at 21°C and washed with PBS. EdU incorporation was detected using the Click iT EdU proliferation kit with Alexa Fluor 647 dye (ThermoFisher, #C10340) according to the manufacturer’s instructions, and nuclei identified by incubation with Hoechst 33342 (ThermoFisher; #C10340; 1:1000) for 15 minutes at 21°C. Hoechst 33342^+^ nuclei were identified in each image and scored as EdU^+^ when EdU fluorescence was also in the nucleus. The percentage (%) of iPericytes that had incorporated EdU was calculated as (EdU^+^ Hoechst 33342^+^ nuclei / total Hoechst 33342^+^ nuclei) x 100. EdU assays were performed on iPericytes cultured in a standard incubator, on myelin-coated plates, under hypoxic conditions (0.5% O_2_), on myelin coated plates in hypoxic conditions or with TNFα and/or IFNγ treatment.

#### Motility assay

5,000 pericytes were plated per well in 96-well plates coated with poly-L-lysine ± pHrodo conjugated myelin (3µg/cm^2^). Pericytes were incubated for 24 hours before being transferred to a Nikon Eclipse Ti2 microscope in a cage incubator (Okolab) maintained at 5% CO_2_ and 37°C. To assess motility under hypoxic conditions, pericytes were placed in the cage incubator maintained at 1% O_2_, 5% CO_2_ and 37°C for 24 hours prior to imaging. Phase contrast images were collected every 3 minutes for 1 hour. Individual cell nuclei were annotated in each frame using QuPath (v.5.1)^105^. Cell velocity was calculated as the change in XY coordinates of the cell nucleus between frames for each individual cell, divided by the change in time (µm/min). Nucleus displacement was calculated as the change in XY of the nucleus coordinates between the first and last frames (1 hour).

#### Morphology assay

Pericytes were plated into 96-well plates at 5,000 cells per well and cultured for 2 days prior to fixation with 4% paraformaldehyde in PBS for 15 minutes at 21°C, and washed with PBS. Phase contrast images were taken on a Nikon Eclipse Ti2 microscope and the outer edge of each pericyte was traced using QuPath (v.5.1) and the annotation tool. Each pericyte was manually classified using the morphological subtypes described by Brown et al. (2023). Cell area, cell perimeter and max diameter were extracted from each pericyte tracing using the Calculate Features tool. Morphological assessments were performed on iPericytes cultured in a standard incubator, on myelin-coated plates, under hypoxic conditions (0.5% O_2_), or exposed to TNFα (20ng/mL) and/or IFNγ (40ng/mL).

For pericyte chain analysis, pericytes were stained with Hoechst 33342 prior to imaging. Pericytes were determined to be in a chain arrangement if 3 or more cells lined up end to end and were physically touching.

#### Attachment assay

Pericytes were plated into 96-well plates at 5,000 cells per well and immediately transferred to a Nikon Eclipse Ti2 microscope in a cage incubator (Okolab) maintained at 5% CO_2_ and 37°C. Phase contrase images were collected every 10 minutes for 2 hours, and then every hour for 3 hours. The percentage (%) of pericytes that had attached to the plate was quantified in each video frame.

#### Contractility and relaxation assays

An xCELLigence real-time cell analysis electrical impedance assay was used to quantify pericyte contractility or relaxation, as previously published ^39,53^. Briefly, 10,000 pericytes were plated in each well of an e-plate (ACEA Biosciences, cat.#0546983000) with 150µL of CPM, and left to adhere overnight. For contractility, 50nM of ET-1was added to 10µL of CPM, for each well to be assessed. The e-plate was quickly removed from the incubator, 10µL of CPM alone (diluent) or diluted endothelin-1 (50µM) added to each well, and then placed in the xCELLigence system, maintained at 37°C and 5% CO_2_. For relaxation, 10µL of CPM alone (diluent) or adenosine (10µM) was used, but the E-plate was removed from incubator for 2 minutes prior adding CPM or adenosine to induce a contraction in response to the change in environmental conditions. The e-plate was then placed in the xCELLigence system. Electrical impedance, expressed as cell index, was measured every minute for 2 hours. Normalised cell index was calculated by normalising the raw cell index to the cell index value at the first timepoint. For cytokine contractility experiments, pericytes were plated into e-plates and allowed to attach for 3 hours prior to exchanging medium with cytokine containing medium. Pericytes were then incubated for 20 hours prior to contractility assessment.

For live imaging contractility assessment, pericytes from one Control cell line was plated into 96-well plates at 5,000 cells per well and cultured for 24 hours. Cells were then transferred to a Nikon Eclipse Ti2 microscope in a cage incubator (Okolab) maintained at 5% CO_2_ and 37°C. Pericytes were incubated for 1 hour to ensure stability of pericyte contractile state, minimising the effects of moving the pericytes from the culture incubator to the microscope incubator. While the cells were maintained in the incubator, 10µL of CPM alone (diluent) or diluted endothelin-1 was added to each well. Cells were imaged every minute for an hour and frames 1, 20 and 60 used for morphological analysis. The outer edge of each pericyte was traced using QuPath (v.5.1) using the annotation tool to measure cell area at each timepoint. To assess changes in contractility, cell area measurements at frames 20 and 60 were normalised to each individual cell area at frame 1 and expressed as a percentage.

#### Myelin extraction and pHrodo-bead conjugation

Animal experiments were approved by the University of Tasmania Institutional Biosafety Committee and Animal Ethics Committee (A0028218 and A0028287) and were carried out in accordance with the Australian Code of Practice for the Care and Use of Animals for Scientific Purposes (8^th^ edition). Mice were housed in Optimice^®^ microisolator cages (2-5 per cage, Animal Care Systems, Colorado, USA) maintained at 21 ± 2°C and followed a 12-hour light/dark cycle. Mice had access to standard rodent chow (Barrastoc rat and mouse pellets) and water *ad libitum*.

Crude mouse myelin extraction was performed as previously described ^106^. Briefly, n=2 ∼1 year old C57BL/6 mice (1 male and 1 female) were euthanised by CO_2_ asphyxiation and the brains dissected into a sterile 6cm cell culture dish containing 0.32M sucrose in 1M Tris-HCl. Brain tissue was chopped into ∼0.5mm^3^ pieces and transferred to a sterile hand-held rotary homogeniser containing ∼10mL of 0.32M sucrose in 1M Tris-HCl. Brain tissue was homogenised and diluted into 0.32M sucrose in 1M Tris-HCl. The tissue homogenate was then placed on top of 0.83M sucrose in 1M TrisHCl solution in polypropylene ultracentrifuge tubes (Beckman Coutler, cat.#331372). The tissue was centrifuged at 100,000g for 45 minutes at 4°C, so that the myelin debris accumulated at the interface of the two sucrose concentrations. Myelin debris was collected, placed in new ultracentrifuge tubes and centrifuged again. The resulting myelin pellet was washed by resuspension in 1M Tris-HCl buffer, centrifugation, resuspension in sterile PBS, centrifugation before being resuspended again in PBS and stored at −80°C.

Myelin was ultrasonicated using a Bioruptor^®^ Plus sonication device (20x, 20 seconds on, 20 seconds off) in 1.5 mL Bioruptor^®^ Plus TPX microtubes (Diagenode, cat.#C30010010-300). The myelin was then resuspended in 0.1M sodium bicarbonate buffer (pH 8.4), prior to adding 10mM of pHrodo iFL Red STP ester, amine reactive dye (ThermoFisher Scientific, cat.#P36011) in DMSO at a concentration of 0.1µL per µg of myelin for 1 hour at 21°C in the dark. Labelled myelin was centrifuged at 22,000g for 5 minutes at 4°C and resuspended in sterile PBS to wash away excess dye. Finally, the labelled myelin was centrifuged at 22,000g for 5 minutes at 4°C, resuspended in sterile PBS at a concentration of 2µg per µL, and sonicated (10x, 20 seconds on, 20 seconds off), before storage at −20°C for use in experiments.

#### Myelin phagocytosis assay

96-well plates were incubated with poly-L-lysine (0.001%, Merck, cat.#P4707) ± pHrodo-myelin (3µg/cm^2^) for 2 days before plates were washed with DPBS and the pericytes seeded at 5,000 cells per well. Seeded plates were placed in a cage incubator (Okolab) maintained at 5% CO_2_ and 37°C. Phase contrast and fluorescent images were taken every 20 minutes for 24 hours. The percentage (%) of pericytes, identified by DIC imaging, that had pHrodo^+^ puncta, indicative of myelin phagocytosis, was quantified in each video frame. From images collected at the 24-hour timepoint, the outer edge of individual pericytes was traced using QuPath (v.5.1), and the area containing pHrodo labelling was calculated and expressed as a percentage of the total cell area.

#### Inducing hypoxia

To model hypoxic conditions, iPericytes cultured on regular or myelin coated plates, were incubated in a hypoxia chamber (StemCell Technologies, #27310) with a portable oxygen monitor. A 100mm petri dish containing 5mL of dH_2_O was placed in the bottom of the chamber to maintain humidity. The chamber was flushed with 5% CO_2_ in N_2_ until the oxygen level stabilised at 0.5%. Cells under normoxic conditions were place in an identical but unsealed chamber. The chambers were maintained inside an incubator at 37°C for 20 hours.

#### Cytokine exposure

To model inflammatory MS lesion-like conditions, iPericytes were incubated in cytokine containing media. iPericytes were cultured in TNFα (20ng/mL; StemCell Technologies, #78157.1) and/or IFNγ (40ng/mL; StemCell Technologies, #78141.1) containing CPM for 20 hours unless otherwise specified.

#### Statistical analyses

All quantification was performed blind to treatment condition and MS status. Statistical analyses were performed in R (v.4.4.2). All data unless otherwise stated were analysed by restricted maximum likelihood (REML) mixed models with cell line as a random effect (lme4 package v.1.1-35.1). Multiple comparison post hoc (Tukey method) tests were performed utilising the Kenward-Roger method for degrees of freedom (emmeans R v.1.8.9). Analysis of proportional data was performed using a binomial generalised linear mixed model. Statistical significance was set as *p*<0.05 and data are presented as group means ± SEM for column and line graphs, and median ± interquartile range for boxplots.

## Data Availability

All data produced in the present study are available upon reasonable request to the authors.

## Acknowledgements

This research was supported by grants from the Medical Research Future Fund (EPCD0008), the National Health and Medical Research Council (NHMRC; 2030057; 2035302; APP1163384), MS Australia (118273, 20-137, 23-0202, 25- 0338) and the Irene Phelps Charitable Trust. A.J.F was supported by an Australian Research Training Program scholarship and a Menzies Institute for Medical Research Philanthropic Fellowship. K.M.Y was supported by a Senior Research Fellowship from MS Australia (21-3-023). J.L.F and N.B.B were supported by Research Fellowships from MS Australia (23-PDF-0121, 22-4-097). B.V.T was supported by an NHMRC Investigator Award (GNT3008389). J.M.C was supported by Tasmanian Graduate Research scholarships.

## Author Contributions

A.J.F: methodology; funding acquisition; validation; investigation; formal analysis; visualisation; writing – original draft; writing – review and editing. N.E.K: methodology; validation; investigation; writing – review and editing. A.G: investigation. R.A.M: investigation; formal analysis; writing – review and editing. J.M.C: methodology; investigation; writing – review and editing. K.P.B: methodology; supervision. B.V.T: conceptualisation; funding acquisition. B.A.S: funding acquisition; methodology; supervision; writing – review and editing. J.L.F: conceptualisation; investigation; funding acquisition; methodology; supervision; writing – review and editing. N.B.B: conceptualisation; funding acquisition; methodology; formal analysis; visualisation; supervision; writing – review and editing. K.M.Y: conceptualisation; funding acquisition; methodology; visualisation; supervision; writing – original draft; writing – review and editing.

