## Supplementary Figures for "Multiple sclerosis iPSC derived-pericytes contract poorly but respond robustly to lesion-relevant environmental stimuli"

**Supplementary Figures and Supplementary Tables** for Fortune et al. *Multiple sclerosis iPSC derived-pericytes contract poorly but respond robustly to lesion-relevant environmental stimuli.*
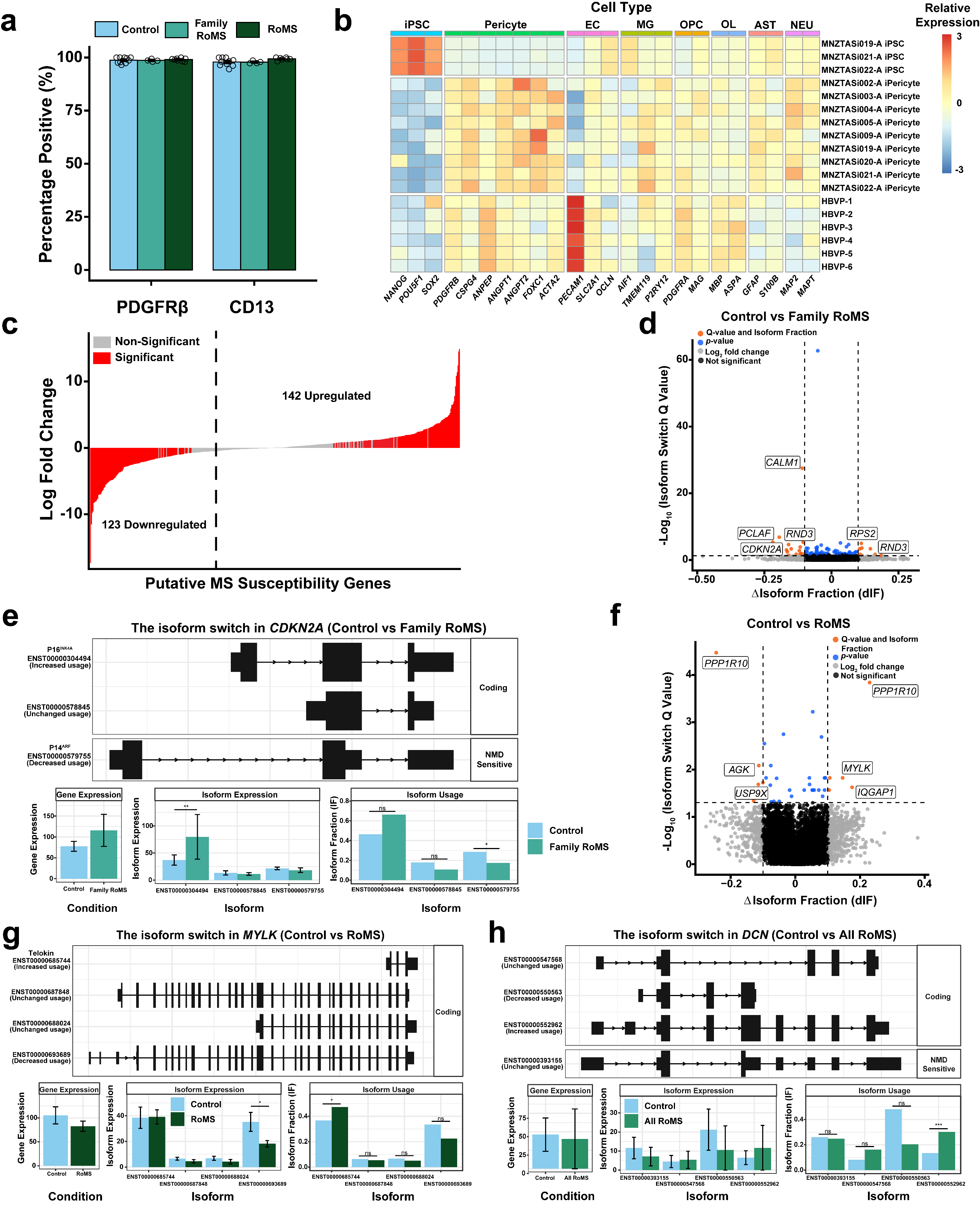
**Figure S1: iPericytes are similar to primary pericytes and exhibit differential transcript usage associated with MS status**

(a) Quantification of the proportion of cells within iPericyte cultures that are PDGFRβ^+^ or CD13^+^. Data is expressed as mean ± SEM and was analysed using a binomial generalised linear mixed model. 1 well stained per cell line for n=5 Control, n=4 Family RoMS and n=8 RoMS cell lines (>100 cells evaluated per line).

(b) Heatmap displaying the relative expression of RNA transcripts from genes associated with cell type identity. Samples include human brain vascular pericytes (HVBP), iPSCs, iPericytes (n=3-5 Control and n=4 Family RoMS lines).

(c) Differential gene expression analysis comparing expression of the putative MS risk genes, identified by the International Multiple Sclerosis Genetics Consortium (2019), between iPSCs and iPericytes, regardless of MS status. Red bars represent significant DEGs that are downregulated (left) or upregulated (right) as the cells differentiate into iPericytes. Thresholds were adjusted *p*-value < 0.05 and Log_2_ fold change >0.585

(d) Volcano plot showing genes that have isoform switches between Control and Family RoMS iPericytes (n=10 Control and n=4 Family RoMS).

(e) Switch plot displaying change in isoform usage in *CDKN2A* between P16^INK4A^ and P14^ARF^ in Control and Family RoMS iPericytes (n=10 Control and n=4 Family RoMS).

(f) Volcano plot showing genes that have isoform switches between Control and RoMS iPericytes (n=10 Control and n=8 RoMS).

(g) Switch plot displaying change in isoform usage in *MYLK* between ENST00000685744 (telokin) and ENST00000693689 in Control and RoMS iPericytes (n=10 Control and n=8 RoMS).

(h) Switch plot displaying change in isoform usage in *DCN* between ENST00000393155 and ENST00000552962 in Control and RoMS iPericytes (n=10 Control and n=8 RoMS).

EC – Endothelial cell, MG – Microglia, OPC – Oligodendrocyte precursor cell, OL – Oligodendrocyte, AST – Astrocyte, NEU – Neuron.

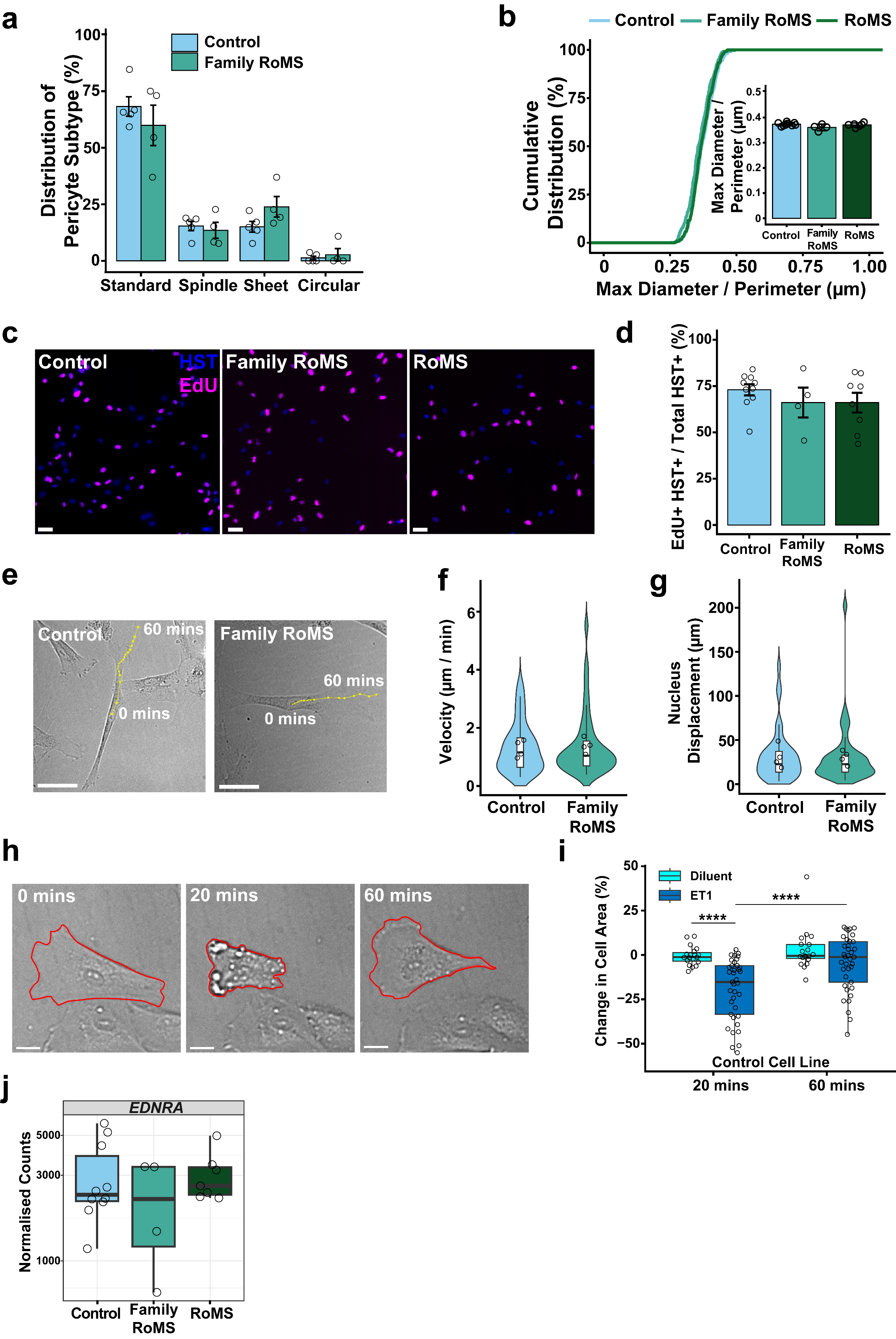

### **Figure S2: MS status does not alter iPSC-derived pericyte proliferation, motility or attachment to culture plates**

(a) The percentage of Control and Family RoMS iPericytes per culture that adopt the standard, spindle, sheet or circular morphological subtype. > 22 cells analysed per culture for n=5 Control and n=4 Family RoMS lines. Data are expressed as mean ± SEM, with dots representing the cell line mean.

(b) Cumulative distribution plot of max feret diameter (max diameter/perimeter) for iPericytes in Control, Family RoMS and RoMS cultures. > 50 cells were evaluated from n=10 Control, n=4 Family RoMS and n=8 RoMS lines. Inset graph represents the mean ± standard deviation for max feret diameter per iPSC line (individual data points represent the mean for each cell line).

(c) Representative images (10x) of iPericytes that incorporated EdU (magenta) over 20 hours. Nuclei are visible due to Hoechst 33342 staining (blue). Scale bars, 50µm.

(d) Quantification of the percentage of Control, Family RoMS and RoMS iPericytes that incorporated EdU over a 20 hour period. Data are presented as mean ± SEM. Individual data points represent the average of 4-9 wells per cell line for n=10 Control, n=4 Family RoMS and n=8 RoMS lines.

(e) Representative images of Control and Family RoMS iPericytes, showing their position at time (t) = 0 mins of the motility assay. The yellow line indicates the direction of iPericyte movement, and each dot represents the location of the nucleus at the end of each 3-minute interval, across the 1 hour imaging period. Scale bars, 50µm.

(f) The mean velocity of iPericytes in Control and Family RoMS cultures. Velocity was calculated by dividing the distance moved between intervals with interval length. > 10 cells were evaluated per n=4 Control and n=4 Family RoMS cell lines. Data are expressed as median ± interquartile range, dots represent cell line mean.

(g) The mean nucleus displacement for iPericytes in Control and Family RoMS cultures. Mean nucleus displacement represents the distance between the nucleus position at t = 0 and t = 60 minutes. > 10 cells were evaluated for n=4 Control and n=4 Family RoMS lines. Data are expressed as median ± interquartile range, and dots represent the mean data for individual cell lines.

(h) To confirm the ability of endothelin-1 to cause pericyte contraction, time-lapse imaging (20x) of an individual Control iPericyte was completed. Between t=0- and 20, pericytes contracted and then returned to baseline after 60-minutes. Red outline denotes the outer diameter of the cell. Scale bars, 10µm.

(i) Quantification of the change in area of individual Control iPericytes between t=0 and t=20 minutes or t=0 and t=60-minutes following exposure to diluent or endothelin-1. This analysis was performed on iPericytes from an individual line.

(j) Boxplot showing *EDNRA* expression from Control, Family RoMS and RoMS iPericytes. Data are expressed as median ± interquartile range, and individual dot points represent cell line means (n=10 Control, n=4 Family RoMS and n=8 RoMS cell lines).

Restricted maximum likelihood linear mixed model with pairwise comparisons using Tukey’s adjustment for p-values. ^∗∗∗∗^*p* < 0.0001.

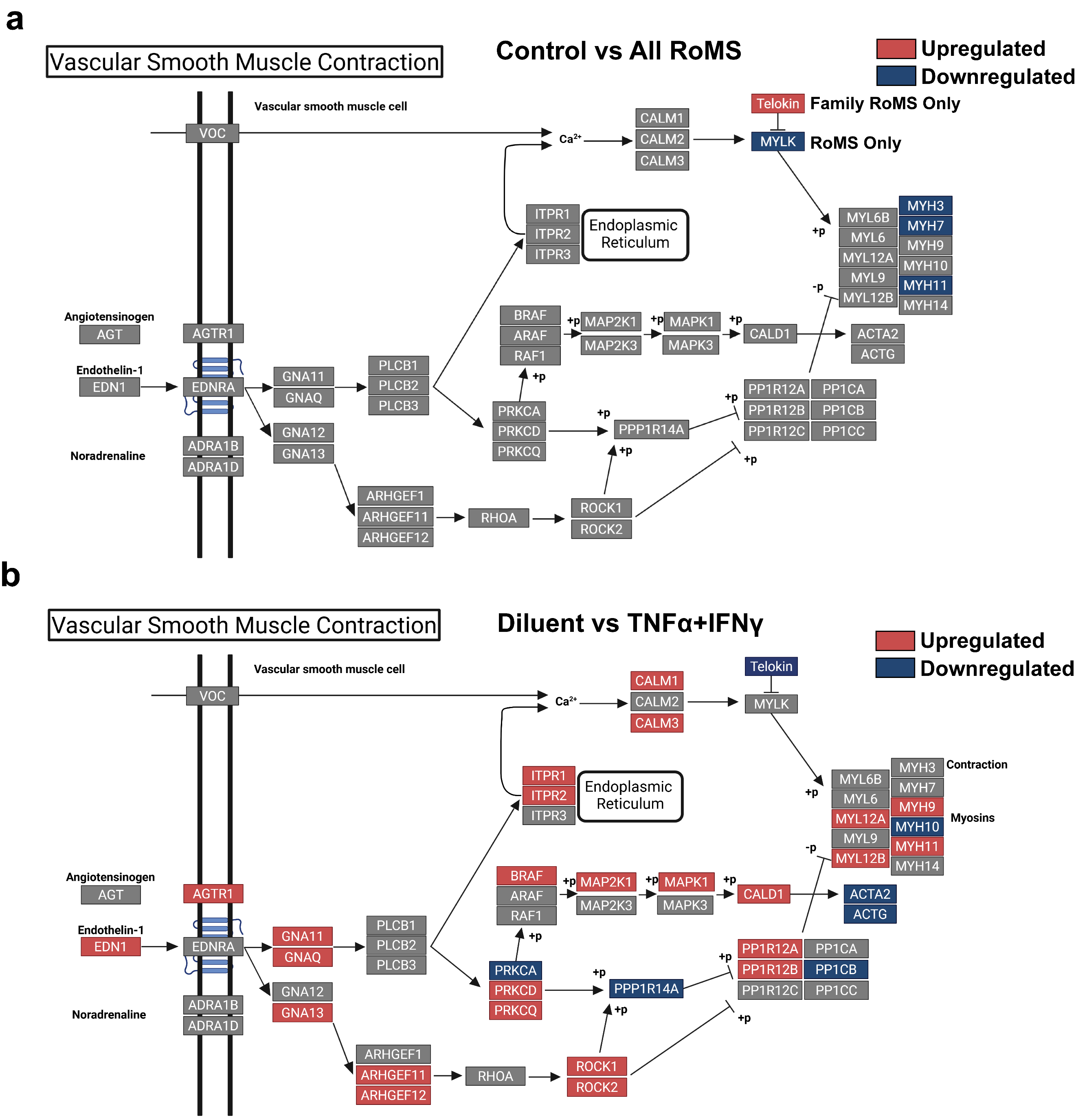

**Figure S3: Changes in expression of genes tagged to the vascular smooth muscle contraction KEGG pathway.**

(a) Genes involved in endothelin-1 and vascular smooth muscle contractile signalling that have altered expression between Control and All RoMS iPericytes. Upregulated genes are in red, downregulated genes are in blue.

(b) Genes involved in endothelin-1 and vascular smooth muscle contractile signalling that have altered expression between diluent and TNFα+IFNγ treated iPericytes. Upregulated genes are in red, downregulated genes are in blue.

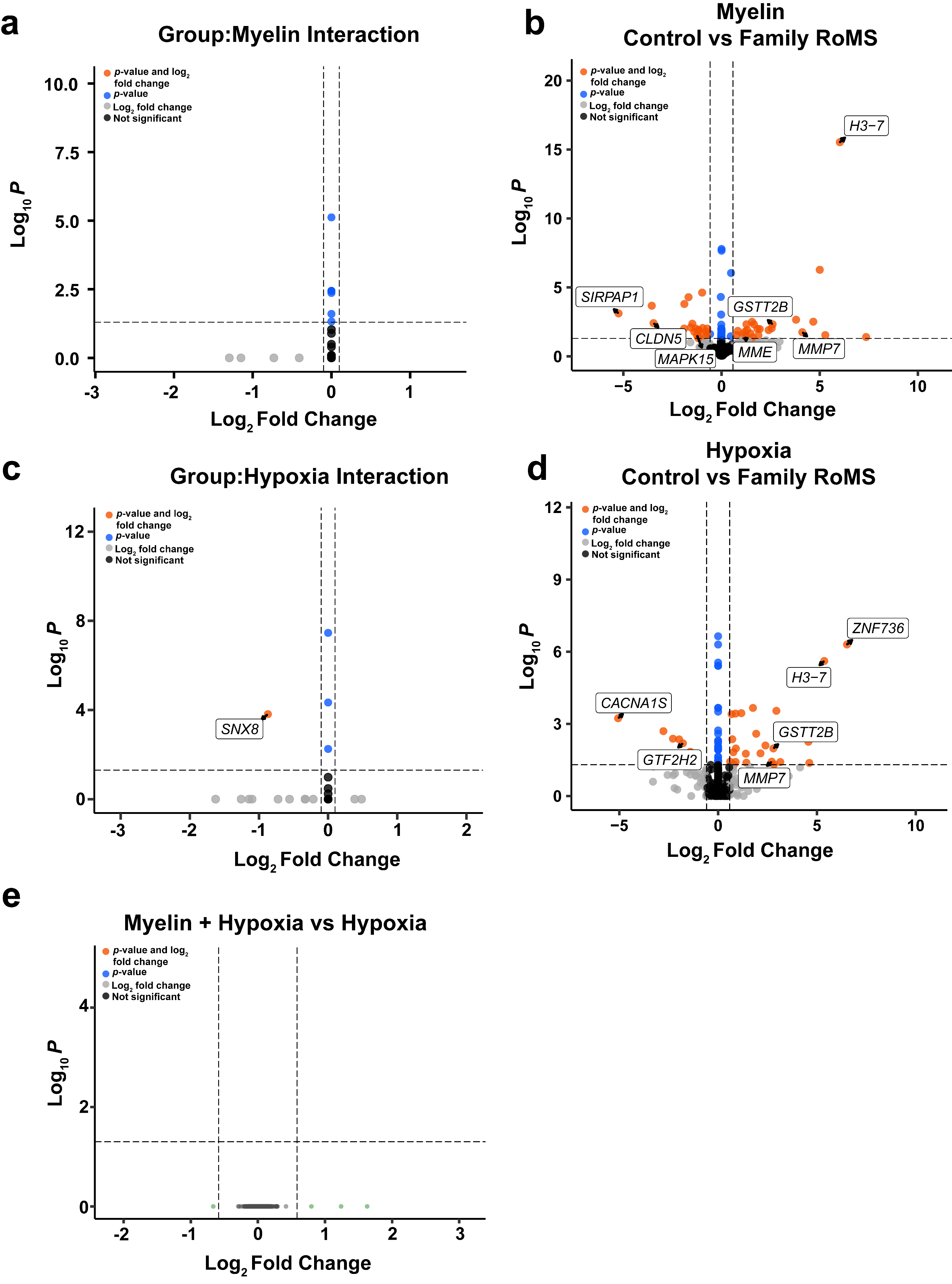

**Figure S4: The transcriptional response of iPericytes to myelin debris or hypoxia is unaffected by MS status, and myelin does not alter hypoxic response.**

(a) Volcano plot of differentially expressed genes with an interaction effect between myelin treatment and disease status. Adjusted *p*-value < 0.05 and Log_2_ fold change ≥ 0.1; n=9 (n=5 Control and n=4 Family RoMS) cell lines.

(b) Volcano plot of differentially expressed genes between Control and Family RoMS iPericytes treated with myelin. The differentially expressed genes identified between myelin treated Control and Family RoMS iPericytes resembled that of the untreated baseline Control and Family RoMS comparison. Adjusted *p*-value < 0.05 and Log_2_ fold change ≥ 0.1; n=9 (n=5 Control and n=4 Family RoMS) cell lines.

(c) Volcano plot of differentially expressed genes with an interaction effect between hypoxia treatment and disease status. Adjusted *p*-value < 0.05 and Log_2_ fold change ≥ 0.1; n=8 (n=5 Control and n=3 Family RoMS) cell lines. Sorting nexin 8 (*SNX8)*, is selectively downregulated in hypoxic Family RoMS iPericytes (Data S8). *SNX8* encodes a protein involved in autophagy, endosome recycling and lysosome tubulation (Li et al., 2024) suggesting that, under hypoxic conditions, Family RoMS iPericytes may have an impaired ability to degrade and remove cellular waste.

(d) Volcano plot of differentially expressed genes between Control and Family RoMS iPericytes exposed to hypoxia. Adjusted *p*-value < 0.05 and Log_2_ fold change ≥ 0.1; n=9 (n=5 Control and n=3 Family RoMS) cell lines.

(e) Volcano plot of differentially expressed genes between myelin+hypoxia or myelin treatment. Adjusted *p*-value < 0.05 and Log_2_ fold change ≥ 0.585; n=8-9 (n=5 Control and n=3-4 Family RoMS) cell lines.

False discovery rate adjustment was used for multiple testing.

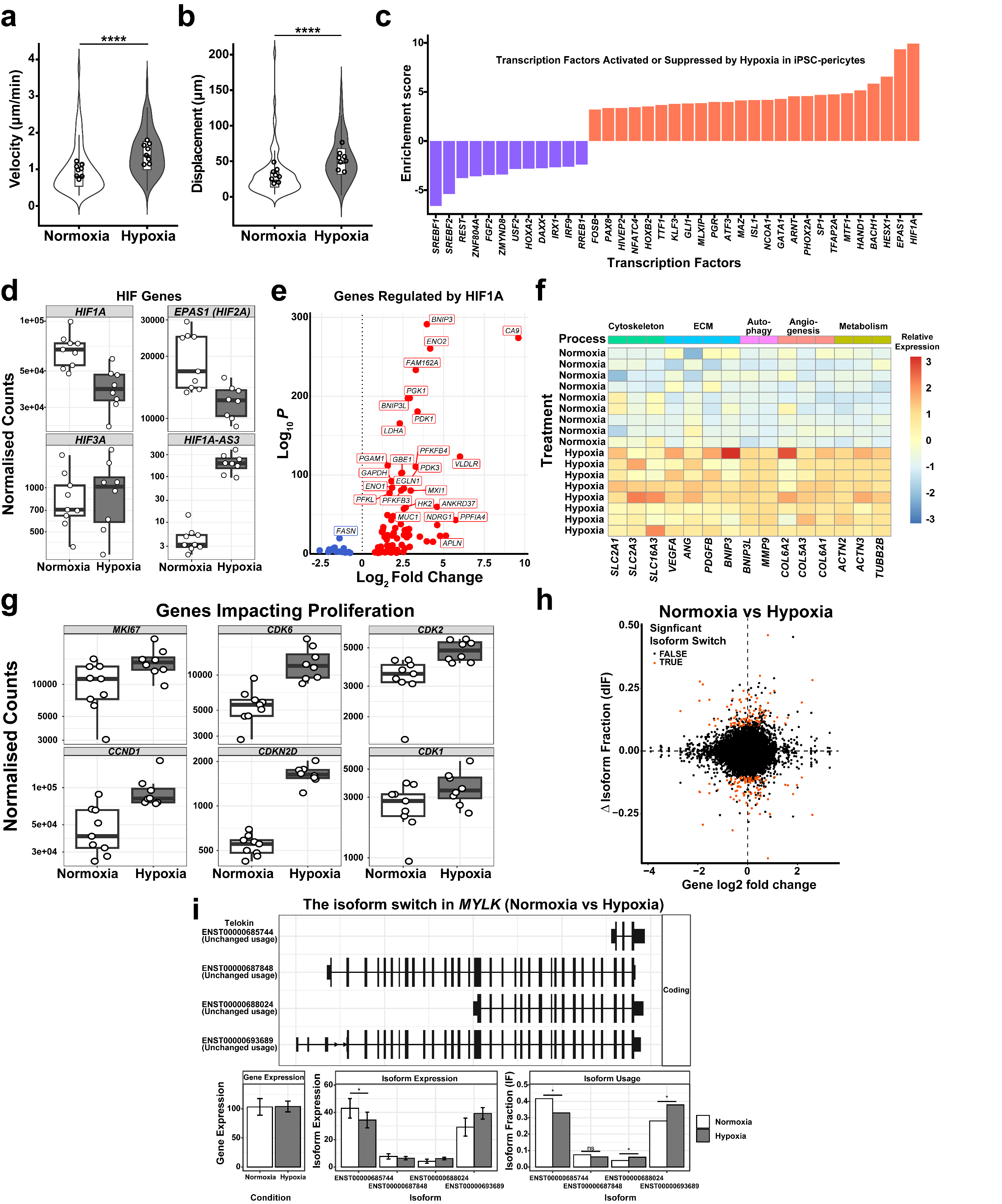

### **Figure S5: Hypoxia modulates motility, proliferation, isoform usage, and HIF-regulated transcription in iPericytes.**

(a) Quantification of the mean velocity for iPericytes cultured under normoxic or hypoxic conditions. Velocity was calculated for > 10 cells per evaluated cell line and was calculated as the distance moved between each interval (3 minutes), divided by the time interval (60 minutes). Data are expressed as median ± interquartile range. As there was no significant interaction between hypoxia and MS status, dots represent the mean for each of the n=8 iPericyte lines analysed (n=4 Control and n=4 Family RoMS).

(b) Quantification of mean nucleus displacement over a 1-hour period for iPericytes cultured under normoxic or hypoxic conditions. > 10 cells were evaluated in each of n=8 (n=4 Control and n=4 Family RoMS) lines. Data expressed as median ± interquartile range and dots represent mean per cell line.

(c) Analysis of RNA sequencing data for iPericytes cultured under normoxic and hypoxic conditions, using decoupleR, revealed an enrichment of differentially expressed genes regulated by particular transcription factors. Transcription factor enrichment scores represented by red bars are upregulated networks, while transcription factor enrichment scores represented by blue bars are downregulated networks.

(d) Boxplots of hypoxia inducible factor (HIF) gene expression by iPericytes cultured under normoxic or hypoxic conditions for 20 hours. Data expressed as median ± interquartile range, and individual dot points represent the mean for n=8 cell lines (n=5 Control and n=3 Family RoMS).

(e) Volcano plot of differentially expressed genes that are downstream of HIF1A in the HIF1A transcriptional network. Adjusted *p*-value < 0.05 and Log_2_ fold change ≥ 0.585; n=8 (n=5 Control and n=3 Family RoMS per treatment group) cell lines.

(f) Heatmap of selected HIF1A-responsive genes that are upregulated in iPericytes cultured under hypoxic conditions. These genes are associated with metabolism, angiogenesis, autophagy, extracellular matrix (ECM) regulation, or cytoskeletal regulation, and are regulated by HIF1A.

(g) Boxplots showing the expression of selected cell cycle regulatory genes in iPericytes cultured under normoxic or hypoxic conditions for 20 hours. Gene expression is shown for *MKI67*, *CDK6*, *CDK2*, *CCND1*, *CDKN2D* and *CDK1*. Data are expressed as median ± interquartile range, and individual dots represent the expression for n=8 cell lines (n=5 Control and n=3 Family RoMS).

(h) Scatter plot displaying the isoform fraction and log fold change in expression for each gene comparing normoxic and hypoxic iPericytes. n=5 Control and n=3 Family RoMS.

(i) Switch plot displaying change in isoform usage in *MYLK* between ENST00000685744 (telokin) and ENST00000693689 in normoxic and hypoxic iPericytes. n=8 iPericyte lines analysed (n=5 Control and n=3 Family RoMS).

Restricted maximum likelihood linear mixed model with pairwise comparisons using Tukey’s adjustment for p-values. ^∗∗∗∗^*p* < 0.0001. False discovery rate adjustment was used for multiple testing. ECM – extracellular matrix

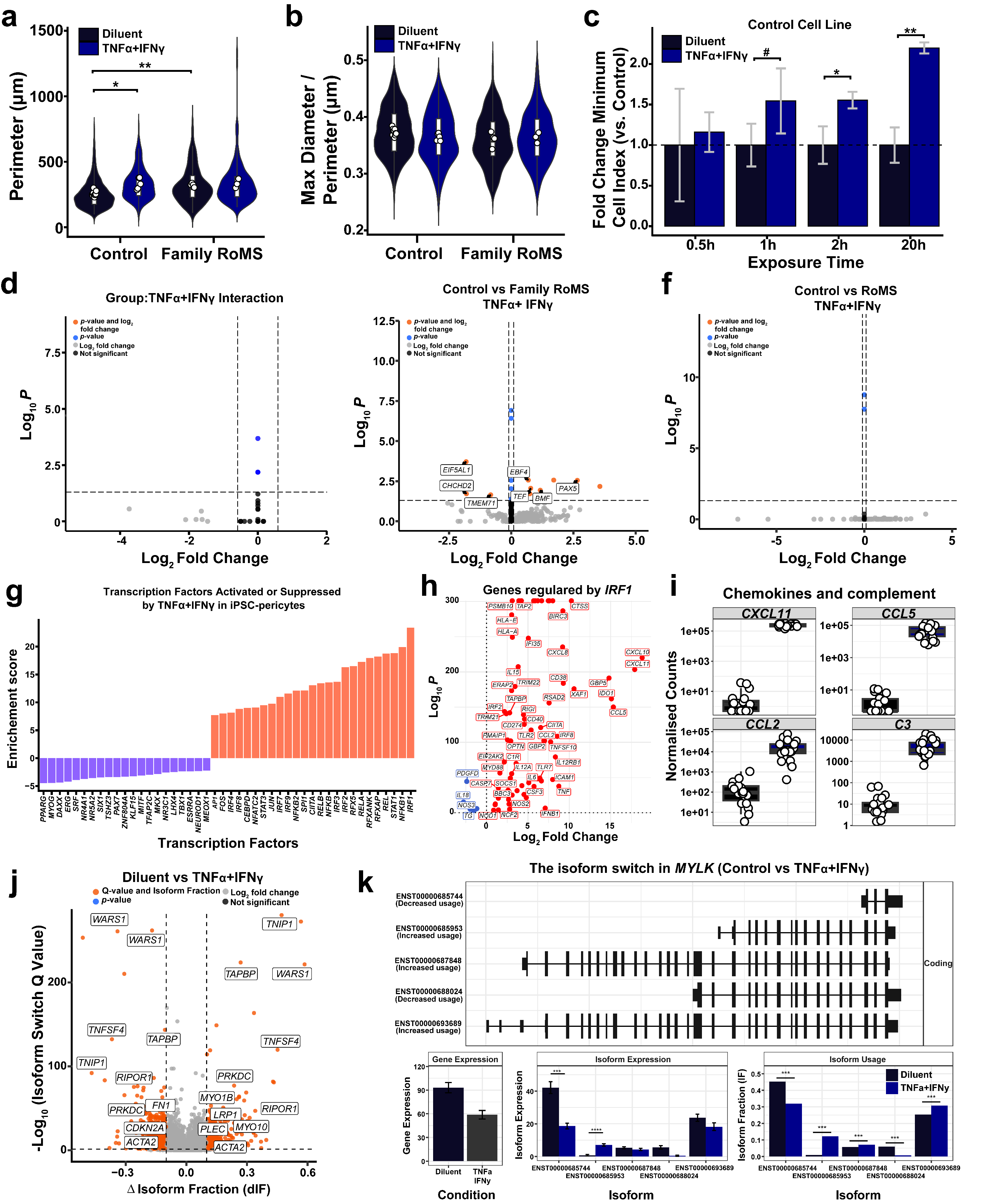

### **Figure S6: TNFα+IFNγ increases iPericyte contractile response to endothelin 1 and alters gene and isoform expression.**

(a) Quantification of Control and Family RoMS iPericyte perimeter following diluent or TNFα+IFNγ treatment. > 50 cells were measured per cell line and treatment condition. Data are expressed as median ± interquartile range, and individual dot points represent the mean for each cell line (n=5 Control and n=4 Family RoMS lines).

(b) Quantification of Control and Family RoMS iPericyte max feret diameter (max diameter/perimeter) following diluent or TNFα+IFNγ treatment. > 50 cells were measured per cell line and treatment condition. Data are expressed as median ± interquartile range, and individual dot points represent the mean for each cell line (n=5 Control and n=4 Family RoMS lines).

(c) iPericytes were exposed to diluent or TNFα+IFNγ for 0.5, 1, 2 or 20 hours. The minimum cell index (maximum contraction) for each well treated with diluent at each timepoint was used to calculate the fold change in contractility due to TNFα+IFNγ. 2 wells per exposure time; n=1 Control cell line.

(d) Volcano plot of differentially expressed genes with an interaction effect between TNFα+IFNγ treatment and RoMS status (all RoMS cases). Adjusted *p*-value < 0.05 and Log_2_ fold change ≥ 0.1; n=8-10 Control, n=4 Family RoMS + n=6-7 RoMS cell lines.

(e) Volcano plot of differentially expressed genes between Control and Family RoMS iPericytes following TNFα+IFNγ treatment. Adjusted *p*-value < 0.05 and Log_2_ fold change ≥ 0.1; n=8 Control and n=4 Family RoMS cell lines.

(f) Volcano plot of differentially expressed genes between Control and RoMS iPericytes following TNFα+IFNγ treatment. Adjusted *p*-value < 0.05 and Log_2_ fold change ≥ 0.1; n=8 Control and n=6 RoMS cell lines.

(g) Analysis of iPericytes cultured with diluent or TNFα+IFNγ conditions reveals an enrichment of differentially expressed genes regulated by particular transcription factors. Transcription factor enrichment scores represented by red bars are upregulated networks, while transcription factor enrichment scores represented by blue bars are downregulated networks.

(h) Volcano plot of differentially expressed genes that are downstream of IRF1 in the IRF1 transcriptional network. Adjusted *p*-value < 0.05 and Log_2_ fold change ≥ 0.5; n= 22 (n=10 Control, n=4 Family RoMS and n=8 RoMS per treatment group) cell lines.

(i) Boxplots showing gene expression from diluent and TNFα+IFNγ treated iPericytes, for differentially expressed genes that encode cytokine and complement proteins. Data are expressed as median ± interquartile range, and individual dot points represent cell line means (n=8-10 Control, n=4 Family RoMS and n=6-7 RoMS cell lines per treatment condition). Data are expressed as mean ± SEM.

(j) Volcano plot showing genes that have isoform switches between diluent and TNFα+IFNγ treated iPericytes (n=8-10 Control, n=4 Family RoMS and n=6-7 RoMS per treatment group).

(k) Switch plot displaying change in isoform usage in *MYLK* between ENST00000685744 (telokin) and ENST00000685953 in diluent and TNFα+IFNγ treated iPericytes (n=8-10 Control, n=4 Family RoMS and n=6-7 RoMS per treatment group).

Restricted maximum likelihood linear mixed model with pairwise comparisons using Tukey’s adjustment for p-values. ^#^ < 0.1, ^∗^*p* < 0.05, ^∗∗^*p* < 0.01.

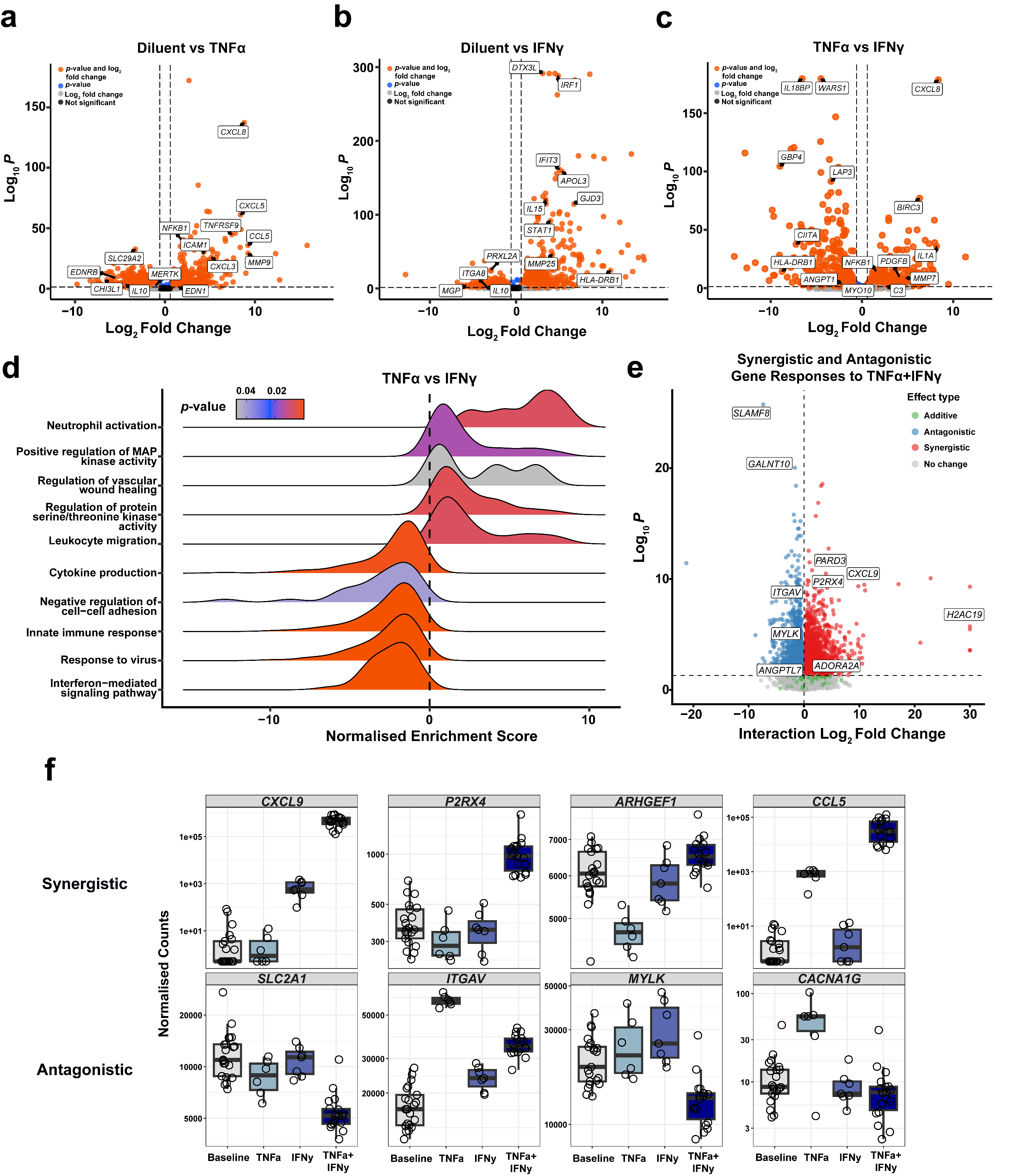

### **Figure S7: TNFα and IFNγ drive differential and interactive gene expression programs in iPericytes.**

(a) Volcano plot of differentially expressed genes between diluent or TNFα treated iPericytes. Adjusted *p*-value < 0.05 and Log_2_ fold change ≥ 0.585; n=9 (n=4 Control and n=4 Family RoMS) cell lines.

(b) Volcano plot of differentially expressed genes between diluent or IFNγ treated iPericytes. Adjusted *p*-value < 0.05 and Log_2_ fold change ≥ 0.585; n=9 (n=4 Control and n=3 Family RoMS) cell lines.

(c) Volcano plot of differentially expressed genes between TNFα or IFNγ treated iPericytes. Adjusted *p*-value < 0.05 and Log_2_ fold change ≥ 0.585; n=9 (n=4 Control and n=3-4 Family RoMS) cell lines.

(d) Ridge plot showing gene set enrichment analysis results comparing TNFα or IFNγ treated iPericytes.

(e) Volcano plot showing the interaction effect between TNFα and IFNγ on gene expression in iPericytes. Genes are coloured by interaction type: synergistic (red), antagonistic (blue), additive (green), or no significant change (grey). Selected genes of interest are labelled.

(f) Boxplots showing the expression of selected genes that exhibit either synergistic or antagonistic TNFα x IFNγ interaction effects in iPericytes. Gene expression is shown for *CXCL9*, *P2XR4*, *ARHGEF1*, *CCL5, SLC2A1*, *ITGAV*, *MYLK* and *CACNA1G*. Data are expressed as median ± interquartile range, and individual dots represent the expression for n=7-22 cell lines (n=5-10 Control, n=3-4 Family RoMS and n=7 RoMS).
